# Continence problems and mental health in adolescents from a UK cohort

**DOI:** 10.1101/2022.12.07.22283198

**Authors:** Katie Gordon, Naomi Warne, Jon Heron, Alexander von Gontard, Carol Joinson

## Abstract

**Background:** Adolescents with continence problems experience a unique range of threats to their psychological wellbeing including perceived stigma, social isolation, and poor self-image. Despite this, the long-term mental health sequelae of adolescent continence problems are unknown.

**Methods:** We used data from the Avon Longitudinal Study of Parents and Children (n=7,332: 3,639 males, 3,693 females) to examine prospective relationships between self-reported incontinence/lower urinary tract symptoms (LUTS) at 14 years (daytime wetting, bedwetting, soiling, urgency, frequent urination, low voided volume, voiding postponement, and nocturia) and common mental health problems at 18 years (any common mental disorder, depression, anxiety, self-harm, and disordered eating). We estimated multivariable regression models adjusted for sex, socioeconomic position, developmental level, IQ, stressful life events, maternal psychopathology, body mass index, and earlier emotional/behavioural problems.

**Findings:** Daytime wetting and voiding postponement showed the greatest number of associations with mental health problems. All incontinence subtypes/LUTS were associated with increased odds of generalised anxiety disorder symptoms [e.g., odds ratio for daytime wetting= 3.01, 95% confidence interval (1.78, 5.09), p<0.001] and/or higher anxiety scores. There was also evidence of associations with common mental disorder [e.g., voiding postponement: 1.88 (1.46, 2.41), p<0.001], depression [e.g., urgency: 1.94 (1.19, 3.14), p=0.008], depressive symptoms [e.g., daytime wetting: 1.70 (1.13, 2.56), p=0.011], self-harm thoughts [e.g., voiding postponement: 1.52 (1.16, 1.99), p=0.003], and disordered eating [e.g., nocturia 1.72 (1.27, 2.34), p=0.001].

**Interpretation:** Incontinence/LUTS at age 14 are associated with increased vulnerability to mental health problems in late adolescence. Treatment of incontinence/LUTS should be integrated with psychological support to mitigate long-term sequelae.

## Introduction

Incontinence and lower urinary tract symptoms (LUTS) in adolescence are poorly understood due to a lack of empirical evidence. Clinicians are often unaware of the wider issues that affect young people with incontinence/LUTS, and this can lead to negative clinical care experiences and inadequate treatment.^1^ It is often believed that incontinence is a problem that resolves during childhood, but persistent, as well as new-onset, cases of incontinence are not uncommon in adolescence.^2,3^ It is estimated that 1–3% of adolescents experience bedwetting, daytime wetting and/or soiling (faecal incontinence).^3^ In-depth qualitative research with 11-17-year-olds with incontinence/LUTS (urinary and/or faecal incontinence, and/or urgency) found that many young people feel hopeless and pessimistic about their prognosis, and they find it challenging to adhere to treatments.^1^ Fears of bullying, social isolation and “feeling like an outsider” are also common, due to the shame and perceived stigma of incontinence.^4^ Young people also report problems in their interpersonal relationships and restrictions to their social life, and feel they need to conceal their continence problems from friends and romantic partners to appear ‘normal’.^4^ Adolescence is a sensitive period for development of self-concept, and peer rejection can lead to negative self-beliefs which increase the risk of mental disorder.^5^ The secondary school environment is particularly challenging for young people with continence problems, with many experiencing anxiety about restricted access to toilets during class, adverse impacts on learning, and disruptions to lessons and exams due to frequent toilet trips.^4^

Cross-sectional associations have been found between urinary incontinence in adolescence and adverse psychosocial outcomes including poor self-image, peer problems, and negative school experiences.^6^ Most mental health problems begin in adolescence, and young people with continence problems could be at greater risk of developing mental disorders due to the unique myriad of stressors they experience in their daily lives. Despite this, the longer-term mental health sequelae of adolescent continence problems are unknown. Evidence-based knowledge is needed to improve support for this vulnerable group.

We examined if incontinence (daytime wetting, bedwetting, soiling) and LUTS (urgency, frequent urination, low voided volume, voiding postponement, nocturia) at age 14 are prospectively associated with mental health problems at age 18. We studied a range of mental health problems that commonly emerge in adolescence including, depression, anxiety, self-harm, and disordered eating.

## Methods

### Participants

Data were obtained from the Avon Longitudinal Study of Parents and Children (ALSPAC) – a large UK-based prospective cohort. ALSPAC recruited pregnant women living in the Avon area of Bristol (UK) with an expected delivery date between 1^st^ April 1991 and 31^st^ December 1992, yielding a cohort of 14,541 pregnancies, 14,676 foetuses, and 13,988 children alive at 12 months.^7,8^ When the oldest study children were 7 years, an attempt was made to increase the sample with participants who failed to enrol during original recruitment. This increased the sample to 15,454 pregnancies, 15,645 foetuses, and 14,865 children alive at 12 months.^9^ The study website contains details of all data that is available through a fully searchable data dictionary and variable search tool (http://www.bristol.ac.uk/alspac/researchers/our-data/). Ethical approval was obtained from the ALSPAC Ethics and Law Committee and the Local Research Ethics Committee. Informed consent for use of data collected via questionnaires and clinics was obtained from participants following the recommendations of the ALSPAC Ethics and Law Committee at the time.

### Continence problems at age 14

Participants completed a questionnaire with items on the frequency of incontinence and LUTS during the previous two weeks including: daytime wetting, bedwetting, and soiling; symptoms of urgency (sudden need to urinate); frequent urination (>7 times per day); low voided volume (passing small amounts of urine); voiding postponement (consciously deferring micturition), and nocturia (waking at night to urinate). Supplementary Table S1 indicates the questions and coding for the continence problem exposures.

### Mental health problems at age 18

#### Depression and depressive symptoms

A computerised version of the Clinical Interview Schedule (CIS-R)^10^ was completed by young people at an assessment clinic they attended at mean age 17.8 years (SD = 0.42), hereafter referred to as 18 years. The CIS-R measures affective and anxiety disorders in the past week and enables diagnoses from the International Statistical Classification of Diseases, 10th Revision (ICD-10) for common mental disorders (CMD) (a score of ≥12 is used to define CMD cases). The depression diagnosis examined in the current study was any depressive episode (mild, moderate or severe). Depressive symptoms were assessed using the Short Mood and Feelings Questionnaire (SMFQ).^11^ Consistent with previous literature, scores of ≥11 were used to define high levels of depressive symptoms, as this threshold has been shown to have high sensitivity and specificity.^12^

#### Anxiety

Generalised anxiety disorder (presence of GAD symptoms: yes/no) was assessed using the CIS-R.^10^ Participants were additionally asked to complete two subscales (physical and mental concerns) of the Anxiety Sensitivity Index (ASI)^13^; higher ASI scores indicate greater severity of anxiety.

#### Self-harm

Participants attending the research clinic also completed CIS-R questions on acts of self-harm, regardless of suicidal intent, over the past year, and thoughts of self-harm over the past week.^10^ Any acts of self-harm, and any thoughts of self-harm, were recorded as cases, regardless of frequency.

#### Disordered eating

Disordered eating (DE) behaviours at age 18 were assessed via self-completed questionnaire (Youth Risk Behavior Surveillance System).^14^ The questionnaire includes items on compensatory behaviours used to control weight, including excessive exercise (that frequently interfered with daily life or resulted in guilt when missing exercise sessions), fasting (not eating for ≥1-day), purging (vomiting, or taking laxatives or other medicines), as well as binge-eating with loss of control. Our primary DE outcome was any disordered eating (any DE), a composite measure covering any instance of these four behaviours. We also examined the specific DE behaviours and an additional composite measure of any of these behaviours occurring ≥1 per week, consistent with DSM-5 diagnostic criteria (DSM-5 DE). Supplementary Table S2 indicates the exact questions and coding for the mental health outcome variables.

### Confounders

Analyses were adjusted for confounders that were identified based on empirical evidence and in consultation with clinical experts. They included child’s sex, family socioeconomic position (parental occupational social class, maternal educational attainment, family size, ethnicity, home ownership status, material hardship); child’s developmental level and IQ; maternal stressful life events; maternal depression and anxiety; child’s body mass index (BMI), and child’s emotional/behaviour problems. Supplementary Table S3 provides full details of the confounders.

### Statistical analysis

The primary analysis focused on any CMD, ICD-10 depression, high depressive symptoms, GAD symptoms, self-harm acts, and any DE as the outcomes. We used multivariable logistic regression to examine the association between incontinence/LUTS and the mental health outcomes. We conducted secondary analyses that examined additional aspects of the mental health outcomes including self-harm thoughts, physical and mental anxiety scores, specific DE behaviours (excessive exercise, fasting, purging, binge-eating), and any DE behaviour occurring ≥1 per week (DSM-5 DE).

Odds ratios (OR) and regression coefficients (B) were estimated, as appropriate, with reference to the groups without incontinence/LUTS. We used linear regression for the continuous outcomes (ASI scores). Regression models were incrementally adjusted for 1) sex, 2) socioeconomic position, 3) developmental level and IQ, 4) maternal stressful life events and maternal psychopathology (depression and anxiety), and 5) child BMI and earlier emotional/behaviour problems. Analyses were performed using Stata version 16.^15^

### Missing data

The primary analysis focused on an imputed sample of 7,332 individuals. Analyses were also conducted on two complete case samples: 1,528 participants who provided all data related to CMD, depression, anxiety, and self-harm, and 1,375 participants who provided all DE outcomes (Figure 1).

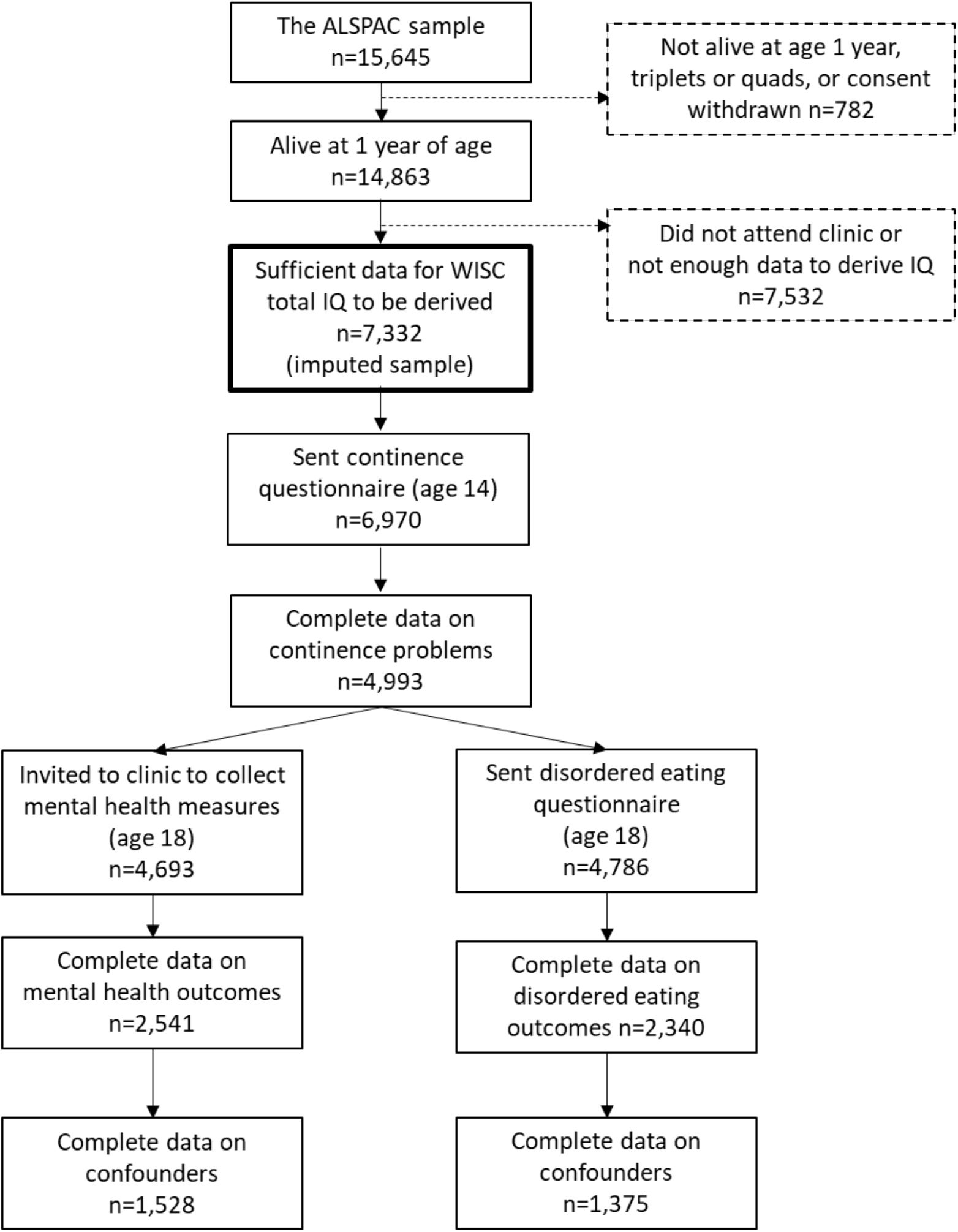
Sample derivation and attrition in ALSPAC ALSPAC – Avon Longitudinal Study of Parents and Children; WISC – Wechsler Intelligence Scale for Children

The amount of missing data for each variable is summarised in Supplementary Table S4. Analyses were conducted on an imputed dataset of 7,332 individuals (3,639 males, 3,693 females) with complete data on IQ (confounder). We restricted the sample to those with complete data on IQ due to a lack of good auxiliary data for IQ, but availability of good auxiliary data for other variables. Missing data on exposures, outcomes and confounders were imputed using the multivariate imputation by chained equations approach (mi impute chained command in Stata) under the Missing at Random (MAR) assumption. In addition to variables used in the main analyses, we included auxiliary variables that were likely to be related to the missing data mechanism including incontinence/LUTS (7 and 9 years), depressive symptoms (10 years), depression diagnosis (15 years), emotional problems (7 years), emotional disorder (15 years), anxiety diagnosis (7 years), behaviour/emotional problems (7 and 9 years), self-harm (16 years), suicidal behaviour (11 years), maternal self-harm (during pregnancy), disordered eating (14 and 16 years), and BMI (8, 10, 12.5 years). We also included earlier measures of key indicators of socioeconomic position (material hardship during pregnancy, home ownership and maternal stressful life events at 6 years), maternal mental health (depressive and anxiety symptoms at 2 years) and child developmental level (6 months). We imputed 100 datasets (a decision informed by examining the Monte Carlo errors for the estimated parameters).

## Results

### Descriptive results

Table 1 shows the descriptive statistics for incontinence/LUTS, mental health problems, and confounders in the imputed sample compared with the complete case samples. The prevalence of incontinence/LUTS and mental health problems were similar in the imputed and complete case samples. The only notable differences were the higher proportions of voiding postponement and high depressive symptoms in the imputed sample compared with the mental health sample. The imputed sample had a lower proportion of females and higher proportions of low parental social class, low maternal education, and non-homeowners.

**Table 1.** Descriptive information on continence problems, mental health problems, and confounders in the imputed sample compared with the complete case samples

| Variable | Imputed sample (n=7,332) |  | Mental health sample (n=1528) |  | Disordered eating sample (n=1375) |  |
| --- | --- | --- | --- | --- | --- | --- |
|  | % or mean (se) |  | % or mean (n or SD) |  | % or mean (n or SD) |  |
| Daytime wetting | 3.49% | (0.28) | 3.60% | (55) | 3.35% | (46) |
| Bedwetting | 3.04% | (0.26) | 2.55% | (39) | 2.62% | (36) |
| Soiling | 4.90% | (0.31) | 4.52% | (69) | 4.95% | (68) |
| Urgency | 5.76% | (0.36) | 4.12% | (63) | 4.80% | (66) |
| Frequent urination | 3.33% | (0.30) | 2.42% | (37) | 3.20% | (44) |
| Low voided volume | 4.85% | (0.33) | 4.38% | (67) | 4.73% | (65) |
| Voiding postponement | 14.40% | (0.51) | 11.65% | (178) | 13.38% | (184) |
| Nocturia | 9.93% | (0.44) | 8.84% | (135) | 8.51% | (117) |
| Common mental disorder | 16.14% | (0.62) | 18.32% | (280) |  |  |
| ICD-10 depression | 8.54% | (0.46) | 6.87% | (105) |  |  |
| High depressive symptoms | 22.48% | (0.69) | 12.76% | (195) |  |  |
| GAD symptom | 6.21% | (0.40) | 5.10% | (78) |  |  |
| Self-harm act | 9.46% | (0.49) | 8.05% | (123) |  |  |
| Any disordered eating | 32.45% | (0.88) | - | - | 33.16% | (456) |
| Sex (female) | 50.37% |  | 55.82% | (853) | 58.84% | (809) |
| Low parental social class | 15.17% | (0.45) | 9.10% | (139) | 8.65% | (119) |
| Ethnicity (non-white) | 4.07% | (0.25) | 3.14% | (48) | 2.91% | (40) |
| Maternal education |  |  |  |  |  |  |
| (O level) | 34.93% | (0.58) | 33.25% | (508) | 32.22% | (443) |
| Vocational or less | 22.25% | (0.51) | 12.37% | (189) | 11.20% | (154) |
| Home ownership (rented/other) | 9.51% | (0.38) | 5.56% | (85) | 5.24% | (72) |
| Family size (3+ children) | 4.68% | (0.26) | 3.47% | (53) | 3.42% | (47) |
| Material hardship | 1.34 | (0.03) | 1.11 | (2.14) | 1.12 | (2.23) |
| Maternal stressful life events | 3.80 | (0.04) | 3.81 | (2.88) | 3.79 | (2.80) |
| Child IQ | 103.97 | (0.19) | 109.84 | (15.39) | 110.04 | (15.92) |
| Child developmental level | 0.01 | (0.01) | 0.02 | (0.90) | -0.01 | (0.92) |
| Maternal depressive symptoms | 5.72 | (0.07) | 5.16 | (5.01) | 5.23 | (5.10) |
| Maternal anxiety symptoms | 4.00 | (0.05) | 3.71 | (3.31) | 3.83 | (3.42) |
| Child behaviour/emotional problems | 6.64 | (0.06) | 5.75 | (4.57) | 5.65 | (4.41) |
| BMI |  |  |  |  |  |  |
| Overweight | 27.17% | (0.59) | 24.35% | (372) | 22.84% | (314) |
| Underweight | 1.89% | (0.19) | 2.23% | (34) | 1.89% | (26) |

BMI – Body Mass Index; GAD – Generalised Anxiety Disorder; ICD-10 - International Classification of Diseases

Family size and material hardship mean scores were similar across the imputed and complete samples. Child mean IQ was lower in the imputed sample, but developmental level was similar across the samples. The proportions of maternal depression/anxiety and child emotional/behaviour problems were similar across imputed and complete samples. The imputed sample had a higher proportion in the overweight category.

Descriptive statistics for the secondary outcomes are shown in Table S5. The proportions of self-harm thoughts, binge-eating and DSM-5 disordered eating were higher in the imputed sample, but the proportions of the other mental health outcomes were similar across the samples.

### Associations between incontinence/LUTS and mental health outcomes

Table 2 shows the unadjusted and fully adjusted results for the primary analysis (based on the imputed data) examining the associations between incontinence/LUTS and mental health outcomes (results for incrementally adjusted models are presented in Table S6). Daytime wetting, urgency, and voiding postponement at age 14 were associated with an increase in the odds of mental health outcomes at age 18 including: CMD, ICD-10 depression, high depressive symptoms (daytime wetting and voiding postponement, but not urgency); the strongest associations were found for presence of GAD symptoms. Most of these associations remained in the fully adjusted models. For example, young people with daytime wetting at age 14 had over a threefold (95% CI: 78% to 409%) increase in the odds of having at least one GAD symptom at age 18 compared to young people without daytime wetting.

**Table 2.** Odds ratios and 95% confidence intervals for the association between continence problems at age 14 and mental health problems at age 18 (results based on imputed sample, n=7,332)

| Exposure | Outcome | Unadjusted |  | Fully adjusted model |  |
| --- | --- | --- | --- | --- | --- |
|  |  | OR (95% CI) | p | OR (95% CI) | p |
| Daytime wetting | Common Mental Disorder | 2.38 (1.59, 3.57) | <.001 | 1.59 (1.03, 2.47) | 0.036 |
| Bedwetting | Common Mental Disorder | 1.77 (1.06, 2.96) | 0.029 | 1.39 (0.78, 2.47) | 0.259 |
| Soiling | Common Mental Disorder | 1.56 (1.08, 2.26) | 0.019 | 1.25 (0.84, 1.86) | 0.278 |
| Urgency | Common Mental Disorder | 2.07 (1.40, 3.06) | <.001 | 1.54 (1.00, 2.36) | 0.050 |
| Frequent urination | Common Mental Disorder | 1.83 (1.07, 3.10) | 0.027 | 1.41 (0.80, 2.50) | 0.231 |
| Low voided volume | Common Mental Disorder | 1.75 (1.16, 2.65) | 0.008 | 1.45 (0.94, 2.26) | 0.095 |
| Voiding postponement | Common Mental Disorder | 2.06 (1.63, 2.60) | <.001 | 1.88 (1.46, 2.41) | <.001 |
| Nocturia | Common Mental Disorder | 1.44 (1.07, 1.94) | 0.017 | 1.19 (0.86, 1.64) | 0.303 |
| Exposure | Outcome | OR (95% CI) | p | OR (95% CI) | p |
| Daytime wetting | ICD-10 depression | 2.48 (1.52, 4.04) | <.001 | 1.77 (1.05, 2.98) | 0.032 |
| Bedwetting | ICD-10 depression | 0.61 (0.22, 1.70) | 0.34 | 0.44 (0.15, 1.31) | 0.140 |
| Soiling | ICD-10 depression | 1.18 (0.67, 2.05) | 0.565 | 0.99 (0.55, 1.78) | 0.964 |
| Urgency | ICD-10 depression | 2.43 (1.56, 3.78) | <.001 | 1.94 (1.19, 3.14) | 0.008 |
| Frequent urination | ICD-10 depression | 1.33 (0.67, 2.67) | 0.413 | 1.04 (0.50, 2.14) | 0.919 |
| Low voided volume | ICD-10 depression | 1.82 (1.10, 2.99) | 0.019 | 1.53 (0.91, 2.57) | 0.107 |
| Voiding postponement | ICD-10 depression | 1.74 (1.29, 2.35) | <.001 | 1.58 (1.15, 2.16) | 0.005 |
| Nocturia | ICD-10 depression | 1.32 (0.89, 1.96) | 0.163 | 1.11 (0.73, 1.69) | 0.628 |
| Exposure | Outcome | OR (95% CI) | p | OR (95% CI) | p |
| Daytime wetting | High depressive symptoms | 2.26 (1.54, 3.32) | <.001 | 1.70 (1.13, 2.56) | 0.011 |
| Bedwetting | High depressive symptoms | 1.42 (0.88, 2.32) | 0.154 | 1.10 (0.63, 1.89) | 0.741 |
| Soiling | High depressive symptoms | 1.83 (1.30, 2.59) | 0.001 | 1.52 (1.05, 2.20) | 0.029 |
| Urgency | High depressive symptoms | 1.27 (0.86, 1.90) | 0.231 | 0.87 (0.56, 1.35) | 0.535 |
| Frequent urination | High depressive symptoms | 1.35 (0.85, 2.13) | 0.202 | 1.01 (0.62, 1.64) | 0.981 |
| Low voided volume | High depressive symptoms | 2.21 (1.55, 3.14) | <.001 | 1.87 (1.28, 2.73) | 0.001 |
| Voiding postponement | High depressive symptoms | 1.68 (1.35, 2.10) | <.001 | 1.53 (1.21, 1.93) | <.001 |

| Nocturia | High depressive symptoms | 1.55 (1.19, 2.03) | 0.001 | 1.26 (0.95, 1.68) | 0.109 |
| --- | --- | --- | --- | --- | --- |
| Exposure | Outcome | OR (95% CI) | p | OR (95% CI) | p |
| Daytime wetting | GAD symptoms | 4.11 (2.51, 6.74) | <.001 | 3.01 (1.78, 5.09) | <.001 |
| Bedwetting | GAD symptoms | 2.50 (1.22, 5.09) | 0.012 | 2.16 (1.03, 4.52) | 0.042 |
| Soiling | GAD symptoms | 1.69 (0.96, 2.96) | 0.067 | 1.32 (0.74, 2.35) | 0.348 |
| Urgency | GAD symptoms | 2.64 (1.54, 4.52) | <.001 | 2.09 (1.19, 3.67) | 0.011 |
| Frequent urination | GAD symptoms | 2.80 (1.48, 5.30) | 0.002 | 2.37 (1.23, 4.57) | 0.010 |
| Low voided volume | GAD symptoms | 1.22 (0.62, 2.42) | 0.564 | 1.03 (0.51, 2.07) | 0.935 |
| Voiding postponement | GAD symptoms | 1.74 (1.22, 2.50) | 0.003 | 1.59 (1.10, 2.30) | 0.013 |
| Nocturia | GAD symptoms | 2.02 (1.31, 3.09) | 0.001 | 1.73 (1.12, 2.68) | 0.014 |
| Exposure | Outcome | OR (95% CI) | p | OR (95% CI) | p |
| Daytime wetting | Self-harm act | 1.66 (0.94, 2.95) | 0.082 | 1.07 (0.58, 1.98) | 0.826 |
| Bedwetting | Self-harm act | 1.98 (1.11, 3.54) | 0.021 | 1.71 (0.93, 3.16) | 0.086 |
| Soiling | Self-harm act | 1.36 (0.83, 2.22) | 0.216 | 1.04 (0.62, 1.73) | 0.885 |
| Urgency | Self-harm act | 0.70 (0.34, 1.45) | 0.337 | 0.49 (0.23, 1.05) | 0.065 |
| Frequent urination | Self-harm act | 1.63 (0.89, 2.98) | 0.112 | 1.32 (0.70, 2.50) | 0.396 |
| Low voided volume | Self-harm act | 0.88 (0.50, 1.55) | 0.645 | 0.71 (0.39, 1.27) | 0.245 |
| Voiding postponement | Self-harm act | 1.17 (0.85, 1.63) | 0.335 | 1.06 (0.75, 1.49) | 0.752 |
| Nocturia | Self-harm act | 1.35 (0.92, 1.97) | 0.126 | 1.14 (0.77, 1.70) | 0.513 |
| Exposure | Outcome | OR (95% CI) | p | OR (95% CI) | p |
| Daytime wetting | Any disordered eating | 2.06 (1.41, 3.01) | <.001 | 1.48 (0.98, 2.25) | 0.064 |
| Bedwetting | Any disordered eating | 1.51 (0.98, 2.30) | 0.059 | 1.61 (1.00, 2.59) | 0.052 |
| Soiling | Any disordered eating | 1.68 (1.21, 2.33) | 0.002 | 1.33 (0.92, 1.91) | 0.131 |
| Urgency | Any disordered eating | 1.33 (0.92, 1.94) | 0.132 | 1.21 (0.79, 1.86) | 0.378 |
| Frequent urination | Any disordered eating | 1.36 (0.90, 2.07) | 0.144 | 1.24 (0.77, 2.00) | 0.368 |
| Low voided volume | Any disordered eating | 1.66 (1.11, 2.48) | 0.014 | 1.52 (0.97, 2.40) | 0.070 |
| Voiding postponement | Any disordered eating | 1.41 (1.13, 1.75) | 0.003 | 1.36 (1.07, 1.74) | 0.013 |
| Nocturia | Any disordered eating | 1.76 (1.34, 2.31) | <.001 | 1.72 (1.27, 2.34) | 0.001 |

Fully adjusted models adjusted for child sex, family socioeconomic position, child's developmental delay and IQ, stressful life events, maternal depression and anxiety, child's BMI and earlier emotional/behaviour problems.
GAD – Generalised Anxiety Disorder; ICD-10 - International Classification of Diseases, 10<sup>th</sup> edition

Bedwetting, frequent urination and nocturia at age 14 were also associated with GAD symptoms at age 18 in the fully adjusted models. Voiding postponement and nocturia were associated with DE, and there was weak evidence for an association with daytime wetting, bedwetting, and low voided volume. There was little evidence of associations between soiling and the mental health outcomes, except for high depressive symptoms. Low voided volume was also associated with high depressive symptoms.

The results for the primary analysis based on the complete data (Table S7) showed evidence of strong associations between daytime wetting and CMD, ICD-10 depression, and GAD symptoms. There were some notable differences between the analyses based on the imputed compared with the complete data, suggesting that the complete case analysis was biased by missing data and/or was underpowered. For instance, there was evidence in the imputed analysis, but not the complete case analysis, that voiding postponement is associated with CMD, ICD-10 depression, GAD symptoms, and DE.

Results for the secondary analysis examining the additional mental health outcomes are presented in Table S8 (imputed data) and Table S9 (complete case analysis). There was evidence in the fully adjusted models for associations with higher physical and/or mental anxiety scores (daytime wetting, soiling, urgency, low voided volume, and voiding postponement) and self-harm thoughts (voiding postponement, daytime wetting, and nocturia). There was also evidence for associations with DSM-5 DE (daytime wetting, soiling, frequent urination, and a weak association with voiding postponement) and DE behaviours including fasting (frequent urination, voiding postponement, and nocturia); purging (daytime wetting); binge-eating (daytime wetting, soiling, low voided volume, nocturia, and a weak association with voiding postponement), and excessive exercise (bedwetting and voiding postponement).

## Discussion

To our knowledge, this is the first prospective cohort study to examine the relationship between incontinence/LUTS and common mental health problems in adolescents. Compared to young people without incontinence/LUTS, adolescents who experienced these problems at age 14 were more likely to have a range of common mental health problems at age 18.

Daytime wetting and voiding postponement were associated with the highest number of mental health problems including CMD, ICD-10 depression, depressive symptoms, and GAD symptoms. Comorbidity has been reported between daytime wetting, voiding postponement and clinically relevant psychological symptoms in children,^16^ but studies of adolescents are lacking.^17^ We found that all types of incontinence/LUTS were associated with increased odds of GAD symptoms and/or higher anxiety scores. Continence problems in children are associated with higher levels of parent-reported anxiety disorders (including GAD), but no studies have specifically focused on adolescence.^18^ We also found associations between incontinence/LUTS and DE behaviours. Inconsistent findings have been reported by studies of small clinical samples that examined if the prevalence of incontinence is greater in adolescents diagnosed with anorexia compared with the general population.^19^ Bedwetting was the least prevalent exposure, which might explain why we found fewer (or only weak evidence of) associations with the mental health problems. We also found few associations between soiling and mental health outcomes. Soiling was defined by a positive response to the question how often do you “Dirty your pants during the day?”. It is possible that some young people responded positively to this question if they had only experienced slightly soiled underwear (rather than an episode of faecal incontinence), which could have resulted in some non-differential misclassification of this exposure.

### Strengths and limitations

Key strengths of this study include the prospective design, the availability of self-reported data on a range of incontinence/LUTS in a large community-based cohort, the use of validated self-report questionnaires for mental health problems, and the availability of data on a wide range of confounders. Data were available on emotional/behaviour problems that preceded the assessment of incontinence/LUTS, thus allowing us to assess whether the presence of pre-existing mental health problems explained our findings. A limitation is that data on DE that clearly preceded incontinence/LUTS were unavailable.

Attrition could have led to selection bias in the complete case analysis because included participants were more socioeconomically advantaged compared with the original cohort. Whilst there is evidence that mental health problems are more common in young people from disadvantaged backgrounds,^20^ the evidence concerning the association between incontinence and socioeconomic background is inconsistent.^21^ We used multiple imputation to address possible bias due to missing data and compared the results from the analysis of the imputed data and the complete case analysis.

The ALSPAC cohort is predominantly white and affluent,^7,8^ hence we are unable to generalise our results to minority ethnic groups and less affluent populations. Further research in these underserved populations is vital to prevent widening inequalities in heath research.

### Interpretation

The mid-teens are a sensitive period for the development of self-image, and there is evidence that continence problems have an adverse impact on a young person’s psychological well-being.^1,4^ Young people with continence problems experience social isolation, perceived stigma, shame, and negative school experiences^4^, all which have been linked to an increased risk of mental health problems.^22,23^ All types of incontinence/LUTS were associated with GAD symptoms and/or higher anxiety scores. Daytime wetting and voiding postponement showed the greatest number of associations with mental health problems. There was also strong evidence that urgency was associated with poorer mental health. Urgency is highly unpredictable in nature, and this could contribute to psychological distress. It is notable that daytime wetting was associated with depression/depressive symptoms and GAD symptoms, whilst bedwetting, possibly due to its low prevalence, showed fewer (and weaker) associations with mental health problems. An alternative explanation is that daytime wetting, compared with bedwetting, is difficult to conceal from peers due to the actions required to manage symptoms (e.g. frequent toilet trips, changing clothes, fear of odour from incontinence pads).^4^ Peer acceptance is strongly valued during adolescence, and qualitative research has found that young people with daytime wetting have a strong desire to hide their problems from peers due to shame and fear of being ostracised.^4^ The perceived stigma of incontinence and difficulty concealing/controlling symptoms might explain why young people with daytime wetting are at increased risk of mental health problems.

Voiding postponement in children is associated with social anxiety, behavioural disorders, daytime wetting and urgency, and is often an acquired and learned behaviour that is used to cope with the perceived embarrassment of needing to use the toilet or to prevent missing out in social situations.^24^ In adolescents, voiding postponement has been linked to fear of using school toilets due to concerns about lack of privacy, hygiene or safety (e.g., bullying) and is associated with an increased risk of LUTS^25.^

The associations between incontinence/LUTS and DE behaviours could be explained by the possibility that incontinence/LUTS and DE could be linked to a need for control and the denial of bodily symptoms.^26^ Most incontinence/LUTS in young people are functional and, consequently, clinicians are often unable to give a medical explanation or specific guidance on treatments and prognosis. This can lead to feelings of uncertainty about the controllability of their continence problem, poor adherence to treatments, and pessimism about future treatment success.^1^ Illness uncertainty has been linked to maladaptive coping, increased psychological distress, depression and reduced quality of life.^27^ Data were unavailable on DE behaviours that clearly preceded incontinence/LUTS, which could lead to the possibility of reverse causality as an alternative explanation for this association. Consequences of DE (e.g., constipation) and behaviours linked to DE (e.g., use of laxatives, fluid restriction, excessive fluid intake)^28^ are also associated with incontinence/LUTS.

Incontinence/LUTS were not associated with self-harm acts, but daytime wetting and voiding postponement were associated with self-harm thoughts. Self-harm acts have been associated with behavioural impulsivity, whereas self-harm thoughts are common among those experiencing affective disorders.^29^ Clinicians should be aware that young people with daytime wetting and voiding postponement are at increased risk of self-harm thoughts, given the association with future suicidal behaviour.^30^

## Conclusion

Incontinence/LUTS in young people have long-term consequences for their mental health, and adolescents with daytime wetting and voiding postponement are particularly vulnerable. Our findings have important clinical implications in terms of highlighting the need for provision of psychological support to minimise the risk of mental health problems in young people with incontinence/LUTS. Despite this, there is a lack of provision of mental health support for these young people. Clinicians who treat incontinence/LUTS often recognise that young people experience psychological distress and have called for mental health support to be routinely available in paediatric continence clinics. There is a gap between paediatric and adult continence services and consequently, adolescents with incontinence/LUTS are an underserved population. Transition from paediatric to adult continence services can be poorly managed, and mental health problems are often not assessed or treated, which could exacerbate existing symptoms and affect treatment adherence. Adult urology services need to know that young people transitioning to adult care are at increased risk of mental health problems. There is also a need for improved support for young people with incontinence/LUTS in secondary schools to manage their symptoms, as well as access to safe, private, and hygienic toilet facilities to prevent young people from avoiding using school toilets. Further research is needed to establish whether the associations we have observed are causal and to identify modifiable factors on the causal pathway from incontinence/LUTS to mental health problems.

## Data Availability

ALSPAC data access is through a system of managed open access. The steps below highlight how to apply for access to the data included in this paper and all other ALSPAC data.
1. Please read the ALSPAC access policy (PDF, 891kB) which describes the process of accessing the data and samples in detail, and outlines the costs associated with doing so.
2. You may also find it useful to browse our fully searchable research proposals database, which lists all research projects that have been approved since April 2011.
3. Please submit your research proposal for consideration by the ALSPAC Executive Committee. You will receive a response within 10 working days to advise you whether your proposal has been approved.
If you have any questions about accessing data or samples, please (data) or (samples).

http://www.bristol.ac.uk/alspac/researchers/access/

## Acknowledgments

We are extremely grateful to all the families who took part in this study, the midwives for their help in recruiting them, and the whole Avon Longitudinal Study of Parents and Children team, which includes interviewers, computer and laboratory technicians, clerical workers, research scientists, volunteers, managers, receptionists, and nurses.

## Data sharing

ALSPAC data access is through a system of managed open access. The steps below highlight how to apply for access to the data included in this paper and all other ALSPAC data.

1. Please read the <u>ALSPAC access policy (PDF, 891kB)</u> which describes the process of accessing the data and samples in detail, and outlines the costs associated with doing so.
2. You may also find it useful to browse our fully searchable <u>research proposals database</u>, which lists all research projects that have been approved since April 2011.
3. Please <u>submit your research proposal</u> for consideration by the ALSPAC Executive Committee. You will receive a response within 10 working days to advise you whether your proposal has been approved.

If you have any questions about accessing data or samples, please (data) or (samples).

