## Supplementary Materials for "Continence problems and mental health in adolescents from a UK cohort"

### Supplementary material

#### Table S1. Derivation of exposure variables used from Avon Longitudinal Study of Parents and Children (ALSPAC)

| Data collection | ALSPAC variable | Question(s)/measure(s) | Responses | Final variable coding |
|---|---|---|---|---|
| **Incontinence** | | | | |
| ALSPAC 'travelling, leisure, and school' questionnaire | | **How often does the following happen to you?** | | |
| | ccp550 | *Daytime wetting*<br>Wet yourself during the day? | 1: Never<br>2: Less than once per week<br>3: Once a week<br>4: 2-5-times a week<br>5: Nearly everyday<br>6: More than once a day<br>-10, -1: Missing | Recode (1=0) (1/6=1)<br>0: No<br>1: Yes |
| | ccp551 | *Bedwetting*<br>Wet the bed at night? | | |
| | ccp552 | *Soiling*<br>Dirty your pants during the day? | | |
| **Lower urinary tract symptoms** | | | | |
| ALSPAC 'travelling, leisure, and school' questionnaire | | **Over the last two weeks, how often have you:** | | |
| | ccp520 | *Urgency*<br>Had the sudden feeling you need a wee and had to dash to the toilet? | 1: Never<br>2: A few times<br>3: Quite often<br>4: A lot<br>-10, -1: Missing | Recode (1,2=0) (3/4=1)<br>0: No<br>1: Yes |
| | ccp521 | *Frequent urination*<br>Had to go to the toilet for a wee more than seven times a day? | | |
| | ccp522 | *Low voided volume*<br>Passed only a small amount when you went for a wee? | | |
| | ccp524 | *Voiding postponement*<br>Avoided going for a wee until the last moment because you were concentrating on other activities? | | |
| | ccp527 | *Nocturia*<br>Woken up to go for a wee? | | |

**Table S2. Derivation of outcome variables used from Avon Longitudinal Study of Parents and Children (ALSPAC)**

| Data collection | ALSPAC Variable | Question(s)/measure(s) | Responses | Final variable coding |
|---|---|---|---|---|
| **Common mental disorders** | | | | |
| Clinical Interview Schedule-Revised[*] (CIS-R)[*] | FJCI050 | Total CIS-R score | Continuous variable (range: 0-39) | Recode (<12=0) (≥12=1)<br>0: No<br>1: Yes |
| **Depression** | | | | |
| Short Moods and Feelings Questionnaire[*] | CCXD917 | High depressive symptoms | Continuous variable (range: 0-26)<br>-10, -1: Missing | Recode (<11=0), (≥11=1)<br>0: No<br>1: Yes |
| CIS-R[*] | FJCI1001 | ICD-10 diagnosis of depression (any severity: mild, moderate, or severe) | 0: Not diagnosed<br>1: Diagnosed<br>-10, -4, -1: Missing | 0: No<br>1: Yes |
| **Anxiety Symptoms** | | | | |
| CIS-R[*] | FJCI602 | Presence of GAD symptoms | Binary variable<br>0: No<br>1:Yes<br>-10, -4, -1: Missing | 0: No<br>1: Yes |
| Anxiety Sensitivity Index[*] | FJLE220 | Physical anxiety subscale score | Continuous variable (range: 10-50)<br>-10, -4, -1: Missing | Continuous variable |
| | FJLE221 | Mental anxiety subscale score | Continuous variable (range: 8-40)<br>-10, -4, -1: Missing | Continuous variable |
| **Self-Harm** | | | | |
| CIS-R[*] | FJCI369 | Have you ever hurt yourself on purpose in any way (e.g., by taking an overdose of pills or by cutting yourself? | 1: No<br>2: Yes<br>-10, -4, -1: Missing | If FJCI369 = 2, recode FJCI370 (1=0) (2/5=1):<br>0: No<br>1: Yes<br>If FJCI369 = 1, recode FJCI370 (.=0) |
| | FJCI370 | How many times have you hurt yourself on purpose in the last year? | 1: None<br>2: Once<br>3: 2-5<br>4: 6-10<br>5: >10<br>-10, -4, -1: Missing | |
| | FJCI371 | During the past week, have you thought about hurting yourself on purpose? | 1: No<br>2: Yes, but never commit suicide<br>3: Yes<br>-10, -4, -1: Missing | Recode (1=0) (1/2=1)<br>0: No<br>1: Yes |
| **Excessive exercise** | | | | |
| ALSPAC 'Your changing life' questionnaire' | | **During the past year:** | | |
| | cct4105 | How often did you do any exercise (going to the gym, brisk walking, or any other sports activity)? | 1: 5 or more times a week<br>2: 1-4 times a week<br>3: 1-3 times a month<br>4: Less than once a month<br>5: Never | **Any excessive exercise:**<br>*Exercise must be accompanied by intent to control weight and/or impairment.*<br><br>Recode cct4105 (5=0) (1/4=1)<br>Recode cct4106 (3=0) (1/2=1)<br>Recode cct4107 (3=0) (1/2=1)<br>Recode cct4108 (0=0) (1/3=1) |
| | cct4106 | Was it difficult for you to do your work or schoolwork because of the amount of time that you were exercising? | 1: Yes, sometimes<br>2: Yes, frequently<br>3: No | |

| Data collection | ALSPAC Variable | Question(s)/measure(s) | Responses | Final variable coding |
|---|---|---|---|---|
| | cct4107 | Did you exercise in order to lose weight or avoid gaining weight? | 1: Yes, sometimes<br>2: Yes, frequently<br>3: No | 0: No (if cct41005, cct4106, cct41007 or cct4108 = 0)<br>1: Yes (if cct41007 = 1 & cct4108 = 1, or if cct41007 =1 & cct4106 = 1)<br><br>**DSM-5† excessive exercise:**<br><br>Recode cct4105 (3/5=0) (1/2=1)<br>Recode cct4106 (3=0) (1/2=1)<br>Recode cct4107 (3=0) (1/2=1), replace = 0 if cct4105 = 0<br>Recode cct4108 (0/1=0) (2/3=1), replace = 0 if cct4105 = 0<br><br>0: (if cct41005, cct4106, cct41007 or cct4108 = 0)<br>1: Yes (if cct41007 = 1 & cct4108 = 1, or if cct41007 =1 & cct4106 = 1) |
| | cct4108 | Did you feel guilty after missing an exercise session? | 0: No<br>1: Yes, sometimes<br>2: Yes, frequently<br>3: Did not miss any exercise sessions | |
| **Fasting** | | | | |
| ALSPAC 'Your changing life' questionnaire' | cct4110 | During the past year, how often did you fast (not eat for at least a day) to lose weight or avoid gaining weight? | 1: Never<br>2: Less than once a month<br>3: 1-3 times a month<br>4: Once a week<br>5: 2 or more times a week | **Any fasting:**<br>Recode (1=0) (2/5=1)<br>0: No<br>1: Yes<br><br>**DSM-5† fasting:**<br>Recode (1/3=0) (4/5=1)<br>0: No<br>1: Yes |
| **Purging** | | | | |
| ALSPAC 'Your changing life' questionnaire' | | **During the past year:** | | |
| | cct4112 | How often did you make yourself throw up (vomit) to lose weight or avoid gaining weight? | 1: Never<br>2: Less than once a month<br>3: 1-3 times a month<br>4: Once a week<br>5: 2-6 times a week<br>6: Every day | **Any purging**<br>Recode cct4112 (1=0) (2/6=1)<br>Recode cct4115 (3=0) (1/2=1)<br>Recode cct4116 (1=0) (2/6=1), replace = 0 if cct4115 = 0<br><br>0: No (if cct4112 & cct4116=0)<br>1: Yes (if cct4112 or cct4116=1)<br><br>**DSM-5† purging**<br>Recode cct4112 (1/3=0) (4/6=1)<br>Recode cct4115 (3=0) (1/2=1)<br>Recode cct4116 (1/3=0) (4/6=1), replace = 0 if cct4115 = 0<br><br>0: No (if cct4112 & cct4116=0)<br>1: Yes (if cct4112 or cct4116=1) |
| | cct4115 | Did you take laxatives or other tablets or medicines (diet pills or water tablets) to lose weight or avoid gaining weight? | 1: Yes, laxative<br>2: Yes, other<br>3: Never | |
| | cct4116 | How often did you take laxatives or other tablets or medicines to lose weight or avoid gaining weight? | 1: Never<br>2: Less than once a month<br>3: 1-3 times a month<br>4: Once a week<br>5: 2-6 times a week<br>6: Every day | |
| **Binge-eating** | | | | |
| | | **During the past year:** | | |

| Data collection | ALSPAC Variable | Question(s)/measure(s) | Responses | Final variable coding |
|---|---|---|---|---|
| ALSPAC 'Your changing life' questionnaire' | cct4120 | During the past year, how often did you go on an eating binge? | 1: Less than once a month<br>2: 1-3 times a month<br>3: Once a week<br>4: More than once a week<br>5: Never | **Any binge-eating**<br>*Binge-eating must be accompanied by loss of control.*<br><br>Recode cct4120 (5=0), (1/4=1)<br>Recode cct4125 (3=0) (1/2=1)<br><br>0: No (if cct4120 or cct4125=0)<br>1: Yes (if cct4112 & cct4125=1)<br><br>**DSM-5† Binge-eating**<br>Recode cct4120 (5=0) (1/2=0) (3/4=1)<br>Recode cct4125 (3=0) (1/2=1)<br><br>0: No (if cct4120 or cct4125=0)<br>1: Yes (if cct4112 & cct4125=1) |
| | cct4125 | Did you feel out of control, like you couldn't stop eating even if you wanted to stop? | 1: Yes, usually<br>2: Yes, sometimes<br>3: No | |
| **Any disordered eating at any frequency** | | | | |
| | | | | **Presence of any fasting, purging, binge-eating, or excessive exercise:**<br>0: No<br>1: Yes |
| **Any disordered eating at DSM-5† frequency** | | | | |
| | | | | **Presence of any DSM-5† fasting, DSM-5† purging, DSM-5† binge-eating, or DSM-5† excessive exercise:**<br>0: No<br>1: Yes |

*Completed at Teen Focus 4 Clinic. †Behaviour occurs ≥1 per week. *Abbreviation: ALSPAC, Avon Longitudinal Study of Parents and Children; DSM-5, Diagnostic and Statistical Manual of Mental Disorders, Fifth Edition.*

**Table S3. Derivation of confounder variables used from Avon Longitudinal Study of Parents and Children (ALSPAC)**

| Data collection | ALSPAC Variable | Question(s)/measure(s) | Responses | Final variable coding |
|---|---|---|---|---|
| **Child Sex** | | | | |
| Recorded at birth | kz021 | Collected from fieldworkers visiting maternity units at birth | 1: male<br>2: female<br>-1: not known | Recode:<br>0: male<br>1: female |
| **Parental social class** | | | | |
| 1991 British Office of Population and Census Statistics job codes | c755<br>c765 | Derived variable from questions:<br>• Actual job, occupation, trade or profession<br>• Please tick which of the following apply to you: foreman, manager, supervisor, leading hand, self-employed, none of these<br>• Type of industry or service given (main things done in job) | 1: I<br>2: II<br>3: III (non-manual)<br>4: III (manual)<br>5: IV<br>6: V<br>65: Armed forces<br>-1: missing | Recode (1/3=0) (4/6=1) (else=.)<br><br>Select highest parental social class i.e., if one parent is non-manual then social class=0:<br><br>0: Non-manual: professional, managerial, or skilled professions<br>1: Manual: partly or unskilled occupations |
| **Ethnicity** | | | | |
| | c804 | | -1: Missing<br>1: White<br>2: Non-White | Recode:<br>0: White<br>1: Non-White |
| **Maternal education** | | | | |
| | c645a | Derived variable: mother's highest educational qualification | 1: CSE/none<br>2: Vocational<br>3: O-level<br>4: A-level<br>5: Degree<br>-1: Missing | Recode (4/5=0) (3=1) (1/2=2)<br>0 = A-level or greater<br>1 = O-level<br>2 = CSE, vocational or less |
| **Home ownership** | | | | |
| | q2010 | **Is your home:**<br>a. being bought/mortgaged<br>b. being bought from council<br>c. owned - with no mortgage to pay<br>d. rented from council<br>e. rented from private landlord - furnished<br>f. rented from private landlord - unfurnished<br>g. rented from housing association<br>h. other (please tick & describe) | 0: Being bought/mortgaged<br>1: Being bought from council<br>2: Owned with no mortgage to pay<br>3: Rented from council<br>4: Rented from private landlord furnished<br>5: Rented from private landlord unfurnished<br>6: Rented from housing association<br>7: other | Recode (0/2=0) (4/5=0) (3=1) (6/7=1)<br>0 = Mortgage/owned/privately rented<br>1 = rent/other |
| **Material hardship** | | | | |
| ALSPAC hardship items | | **How difficult at the moment do you find it to afford these items:** | | Recode r9000-r9004 (5=4)<br>20–(r9000+r9001+r9002+r9003 +9004) = summed score (range: 0-15)<br><br>**Mode imputation:** Replace summed score =. with mode score if at least at least one of r9000-9004 answered |
| | r9000 | a) food | 1: Very difficult<br>2: Fairly difficult<br>3: Slightly difficult<br>4: Not difficult<br>5: Paid directly by social security | |
| | r9001 | b) clothing | | |
| | r9002 | c) heating | | |
| | r9003 | d) rent or mortgage | | |
| | r9004 | e) things you need for your children | | |

| Data collection | ALSPAC Variable | Question(s)/measure(s) | Responses | Final variable coding |
|---|---|---|---|---|
| | | | | (Higher scores = more material hardship) |
| **Family size** | | | | |
| | b032 | Derived using parity as proxy. | Continuous variable (range: 0-22) | Recode (0/2=0) (3/22=1)<br>0 = <3 children<br>1 = ≥3 children |
| **Developmental delay at 18-month** | | | | |
| Adapted from Denver Developmental Screening Test (*Pediatrics*. 1992;89(1):91–7) | kd680 | Derived variable: Total ALSPAC development score at 18 months (complete case) | Continuous score<br>-102: Not in correct age range<br>-101: Missing | Continuous score<br>(-101, -102 = missing) |
| **Child IQ** | | | | |
| Adapted from Wechsler Intelligence Scale for Children (3rd ed) | f8ws112 | Derived variable: Total IQ score at 8-years | Continuous score<br>-3: Not enough subtests done<br>-2 Did not start WISC | Continuous score<br>(-3, -2 = missing) |
| **Stressful life events** | | | | |
| ALSPAC 'life events inventory' questionnaire | | **Since the study child's 9th birthday:** | | |
| | r5000 | Respondent's husband/partner died | 0: Relevant text but no box ticked<br>1: Yes, since study child was 9 or 10<br>2: Yes, since child's 11th birthday<br>3: Yes, both when the study child was 10/11 and since 9th birthday<br>4: No, did not happen<br>-10: Not completed<br>-1: No response | Recode r5000-r5044 (4=0) (0/3=1)<br>Create summed score (range: 0-45)<br>**Mode imputation:** Replace summed score =. with mode score if at least at least one of r5000-5044 answered |
| | r5001 | One of respondent's children has died | | |
| | r5002 | Respondent's friend/relative has died | | |
| | r5003 | One of respondent's children has been ill | | |
| | r5004 | Respondent's husband/partner has been ill | | |
| | r5005 | One of respondent's children has been ill | | |
| | r5006 | Respondent's friend/relative has been ill | | |
| | r5007 | Respondent has been admitted to hospital | | |
| | r5008 | Respondent has been in trouble with the law | | |
| | r5009 | Respondent has been divorced | | |
| | r5010 | Respondent's husband/partner didn't want their child | | |
| | r5011 | Respondent has been very ill | | |
| | r5012 | Respondent's husband/partner lost his job | | |
| | r5013 | Respondent's husband/partner had problems at work | | |
| | r5014 | Respondent had problems at work | | |
| | r5015 | Respondent lost their job | | |
| | r5016 | Respondent's husband/partner went away | | |
| | r5017 | Respondent's husband/partner was in trouble | | |
| | r5018 | Respondent separated from husband/partner | | |
| | r5019 | Respondent's income was reduced | | |
| | r5020 | Respondent argued with their husband/partner | | |
| | r5021 | Respondent argued with their family/friends | | |
| | r5022 | Respondent moved house | | |
| | r5023 | Respondent's husband/partner was physically cruel to them | | |
| | r5024 | Respondent became homeless | | |
| | r5025 | Respondent had a major financial problem | | |
| | r5026 | Respondent got married | | |

| Data collection | ALSPAC Variable | Question(s)/measure(s) | Responses | Final variable coding |
|---|---|---|---|---|
| | r5027 | Respondent's husband/partner was physically cruel to their children | | |
| | r5028 | Respondent was physically cruel to their children | | |
| | r5029 | Respondent attempted suicide | | |
| | r5030 | Respondent was convicted of an offence | | |
| | r5031 | Respondent became pregnant | | |
| | r5032 | Respondent started a new job | | |
| | r5033 | Respondent returned to work | | |
| | r5034 | Respondent had a miscarriage | | |
| | r5035 | Respondent had an abortion | | |
| | r5036 | Respondent has taken an examination | | |
| | r5037 | Respondent's husband/partner was emotionally cruel to them | | |
| | r5038 | Respondent's husband/partner has been emotionally cruel to their children | | |
| | r5039 | Respondent has been emotionally cruel to their children | | |
| | r5040 | Respondent's house/car was burgled | | |
| | r5041 | Respondent found a new partner | | |
| | r5042 | One of respondent's child started school | | |
| | r5043 | Respondent's husband/partner started a new job | | |
| | r5044 | Respondent's pet died | | |
| **Maternal depression** | | | | |
| Edinburgh Postnatal Depression Scale when child was 11-years (*Br J Psychiatry J Ment Sci*. 1987;150:782–6) | | **In the past seven days:** | | |
| | r4010 | I have been able to laugh and see the funny side of things | 1: As much as I always could<br>2: Not quite so much now<br>3: Definitely not so much now<br>4: Not at all<br>-10, -1: Missing | Recode r4010, r4011, r4013: (1=0) (2=1) (2=3) (4=3)<br>Recode r4012, r4014-r4019: (4=0), (3=1) (2=2), (1=3)<br>Sum all 10 items to generate continuous score (range:0-30)<br><br>**Mode imputation:** Replace summed score =. with mode score if at least at least one of r4010-r4019 answered |
| | r4011 | I have looked forward with enjoyment to things | 1: As much as I ever did<br>2: Rather less than I used to<br>3: Definitely less that I used to<br>4: Hardly at all<br>-10, -1: missing | |
| | r4012 | I have blamed myself unnecessarily when things when wrong | 1: Yes, most of the time<br>2: Yes, some of the time<br>3: Not very often<br>4: Never<br>-10, -1: Missing | |
| | r4013 | I have been anxious or worried for no good reason | 1: No, not at all<br>2: Hardly ever<br>3: Yes, sometimes<br>4: Yes, often<br>-10, -1: Missing | |
| | r4014 | I have felt scared or panicky for no good reason | 1: Yes, quite a lot<br>2: Yes, sometimes | |

| Data collection | ALSPAC Variable | Question(s)/measure(s) | Responses | Final variable coding |
|---|---|---|---|---|
| | | | 3: No, not much<br>4: No, not at all<br>-10, -1: Missing | |
| | r4015 | Things have been getting on top of me | 1: Yes, most of the time I haven't been able to cope<br>2: Yes, sometimes I haven't been coping<br>3: No, most of the time I have coped<br>4: No, I have been coping as well as ever<br>-10, -1: Missing | |
| | r4016 | I have been so unhappy that I have had difficulty sleeping | 1: Yes, most of the time<br>2: Yes, sometimes<br>3: Not very often<br>4: No, not at all<br>-10, -1: Missing | |
| | r4017 | I have felt sad or miserable | 1: Yes, most of the time<br>2: Yes, sometimes<br>3: Not very often<br>4: No, not at all<br>-10, -1: Missing | |
| | r4018 | I have felt so unhappy that I've been crying | 1: Yes, most of the time<br>2: Yes, quite often<br>3: Only occasionally<br>4: Never<br>-10, -1: Missing | |
| | r4019 | The thought of harming myself has occurred to me | 1: Yes, quite often<br>2: Yes, Sometimes<br>3: Hardly ever<br>4: Never<br>-10, -1: Missing | |
| **Maternal anxiety** | | | | |
| Crown Crisp Experimental Index when child was 11-years (*Br J Med Psychol*. 1988;61(3):255–66) | r4000 | Do you feel upset for no obvious reason? | 1: Very often<br>2: Often<br>3: Not very often<br>4: Never<br>-10, -1: missing | Recode r4000 r4005 r4006 (3=0) (4=0) (1=2) (2=2)<br>Recode r4001 r4002 r4004 r4007 (4=0) (3=1) (1=2) (2=2)<br>Recode r4003 (4=0) (1=2) (2=2) (3=2)<br>Generate total score: sum of recoded r4000-r4007 (range: 0-24)<br><br>**Mode imputation:** Replace summed score =. with mode score if at least at least one of r4007-r4007 answered |
| | r4001 | Have you felt as though you might faint? | | |
| | r4002 | Do you feel uneasy and restless? | | |
| | r4003 | Do you sometimes feel panicky? | | |
| | r4004 | Do you worry a lot? | | |
| | r4005 | Do you feel strung-up inside? | | |
| | r4006 | Do you ever have the feeling you are going to pieces? | | |
| | r4007 | Do you have bad dreams which upset you when you wake up? | | |
| | ku847 | Wet self during the day | | |
| | ku848 | Wet the bed at night | | |
| **Child body mass index** | | | | |
| Clinical measures of height and weight at 13.5-years | fg3134 | Z-score derived using LMS parameters and 1990 British Growth Reference[1,2]. | Continuous score<br>-110, -106, -101: Missing | Recode z-scores in line with WHO thresholds[3]: |

| Data collection | ALSPAC Variable | Question(s)/measure(s) | Responses | Final variable coding |
|---|---|---|---|---|
| | | | | 0: Healthy weight (z-score >-2 to <1)<br>1: Overweight (z-score: ≥1)<br>2: Underweight (z-score: ≤-2) |
| **Earlier emotional and behavioural problems** | | | | |
| Adapted from Strengths and Difficulties Questionnaire (*Eur Child Adolesc Psychiatry*. 1998;7(3):125–30) | kw660b | Derived variable: SDQ total difficulties score (prorated). | Continuous score (range 0-40)<br>-10, -6, -5: Missing | Continuous score<br>Missing = -10, -6, -5 |

*Abbreviations: ALSPAC, Avon Longitudinal Study of Parents and Children; LMS, lambda-mu-sigma; BMI, body mass index.*

**Table S4. Amount of missing information for each variable in the substantive models**

| Variable | N missing (/7332) | % |
|---|---|---|
| Daytime wetting | 2188 | 29.84 |
| Bedtime wetting | 2187 | 29.83 |
| Soiling | 2195 | 29.94 |
| Urgency | 2179 | 29.72 |
| Frequent urination | 2194 | 29.92 |
| Low voided volume | 2210 | 30.14 |
| Voiding postponement | 2195 | 29.94 |
| Nocturia | 2206 | 30.09 |
| Common Mental Disorder | 3537 | 48.24 |
| ICD-10 Depression | 3537 | 48.24 |
| High depressive symptoms (sMFQ) | 3674 | 50.11 |
| GAD symptoms | 3537 | 48.24 |
| Physical anxiety score | 3711 | 50.61 |
| Mental anxiety score | 3593 | 49.00 |
| Self-harm act | 3537 | 48.24 |
| Self-harm thoughts | 3537 | 48.24 |
| Any disordered eating | 4636 | 63.23 |
| Excessive exercise | 4689 | 63.95 |
| Fasting | 4645 | 63.35 |
| Purging | 4648 | 63.39 |
| Binge-eating | 4650 | 63.42 |
| DSM-5 frequency disordered eating | 4635 | 63.22 |
| Sex | 0 | 0.00 |
| Parental social class | 844 | 11.51 |
| Ethnicity | 664 | 9.06 |
| Maternal education | 556 | 7.58 |
| Home ownership | 1159 | 15.81 |
| Material hardship | 1514 | 20.65 |
| Family size/Parity | 550 | 7.50 |
| Developmental level | 2090 | 28.51 |
| IQ | 0 | 0.00 |
| Maternal stressful life events | 1459 | 19.90 |
| Maternal depression | 1463 | 19.95 |
| Maternal anxiety | 1468 | 20.02 |
| BMI | 2055 | 28.03 |
| Earlier behaviour and emotional problems (SDQ) | 1637 | 22.33 |

**Table S5. Descriptive information for secondary outcomes (imputed and complete case samples)**

| | Imputed sample (n=7,332) | | Mental health sample (n=1528) | | Disordered eating sample (n=1375) | |
|---|---|---|---|---|---|---|
| **Variable** | **% or mean (se)** | | **% or mean (n or SD)** | | **% or mean (n or SD)** | |
| Physical anxiety | 25.44 | 0.11 | 25.40 | 7.51 | | |
| Mental anxiety | 21.84 | 0.08 | 21.68 | 4.81 | | |
| Self-harm thoughts | 11.74% | 0.56 | 9.42% | 144 | | |
| Excessive exercise | 21.89% | 0.81 | | | 22.55% | 310 |
| Fasting | 11.49% | 0.69 | | | 10.76% | 148 |
| Purging | 6.08% | 0.44 | | | 5.53% | 76 |
| Binge-eating | 13.42% | 0.72 | | | 11.71% | 161 |
| DSM-5 disordered eating | 11.85% | 0.64 | | | 9.67% | 133 |

DSM-5 - Diagnostic and Statistical Manual of Mental Disorders, Fifth Edition

**Table S6. Sequential adjustments for primary outcomes using imputed data (n=7,332)**

| | | Unadjusted | | Adjusted 1 | | Adjusted 2 | | Adjusted 3 | | Adjusted 4 | | Adjusted 5 | |
|---|---|---|---|---|---|---|---|---|---|---|---|---|---|
| **Exposure** | **Outcome** | **OR (95% CI)** | **p** | **OR (95% CI)** | **p** | **OR (95% CI)** | **p** | **OR (95% CI)** | **p** | **OR (95% CI)** | **p** | **OR (95% CI)** | **p** |
| Daytime wetting | Common mental disorder | 2.38 (1.59, 3.57) | <.001 | 1.99 (1.32, 3.01) | 0.001 | 1.88 (1.23, 2.86) | 0.003 | 1.89 (1.24, 2.89) | 0.003 | 1.79 (1.17, 2.74) | 0.007 | 1.59 (1.03, 2.47) | 0.036 |
| Bedwetting | Common mental disorder | 1.77 (1.06, 2.96) | 0.029 | 1.90 (1.12, 3.22) | 0.017 | 1.67 (0.97, 2.89) | 0.065 | 1.68 (0.97, 2.89) | 0.063 | 1.66 (0.96, 2.88) | 0.071 | 1.39 (0.78, 2.47) | 0.259 |
| Soiling | Common mental disorder | 1.56 (1.08, 2.26) | 0.019 | 1.34 (0.92, 1.97) | 0.128 | 1.32 (0.90, 1.95) | 0.157 | 1.33 (0.90, 1.96) | 0.151 | 1.30 (0.88, 1.92) | 0.192 | 1.25 (0.84, 1.86) | 0.278 |
| Urgency | Common mental disorder | 2.07 (1.40, 3.06) | <.001 | 1.99 (1.33, 2.98) | 0.001 | 1.74 (1.15, 2.64) | 0.009 | 1.75 (1.16, 2.66) | 0.008 | 1.67 (1.10, 2.54) | 0.017 | 1.54 (1.00, 2.36) | 0.05 |
| Frequent urination | Common mental disorder | 1.83 (1.07, 3.10) | 0.027 | 1.74 (1.01, 3.00) | 0.047 | 1.55 (0.89, 2.71) | 0.12 | 1.56 (0.89, 2.72) | 0.116 | 1.53 (0.87, 2.67) | 0.139 | 1.41 (0.80, 2.50) | 0.231 |
| Low voided volume | Common mental disorder | 1.75 (1.16, 2.65) | 0.008 | 1.64 (1.08, 2.49) | 0.021 | 1.56 (1.02, 2.39) | 0.039 | 1.57 (1.02, 2.40) | 0.039 | 1.55 (1.01, 2.38) | 0.047 | 1.45 (0.94, 2.26) | 0.095 |
| Voiding postponement | Common mental disorder | 2.06 (1.63, 2.60) | <.001 | 2.03 (1.60, 2.58) | <.001 | 1.95 (1.52, 2.49) | <.001 | 1.95 (1.52, 2.49) | <.001 | 1.93 (1.51, 2.46) | <.001 | 1.88 (1.46, 2.41) | <.001 |
| Nocturia | Common mental disorder | 1.44 (1.07, 1.94) | 0.017 | 1.41 (1.04, 1.91) | 0.028 | 1.30 (0.95, 1.77) | 0.103 | 1.30 (0.95, 1.78) | 0.101 | 1.28 (0.93, 1.76) | 0.124 | 1.19 (0.86, 1.64) | 0.303 |
| **Exposure** | **Outcome** | **OR (95% CI)** | **p** | **OR (95% CI)** | **p** | **OR (95% CI)** | **p** | **OR (95% CI)** | **p** | **OR (95% CI)** | **p** | **OR (95% CI)** | **p** |
| Daytime wetting | ICD-10 depression | 2.48 (1.52, 4.04) | <.001 | 2.08 (1.27, 3.40) | 0.004 | 1.95 (1.18, 3.24) | 0.01 | 2.00 (1.20, 3.32) | 0.008 | 1.96 (1.18, 3.24) | 0.009 | 1.77 (1.05, 2.98) | 0.032 |
| Bedwetting | ICD-10 depression | 0.61 (0.22, 1.70) | 0.34 | 0.64 (0.23, 1.79) | 0.389 | 0.52 (0.18, 1.50) | 0.224 | 0.52 (0.18, 1.52) | 0.232 | 0.52 (0.18, 1.52) | 0.232 | 0.44 (0.15, 1.31) | 0.14 |
| Soiling | ICD-10 depression | 1.18 (0.67, 2.05) | 0.565 | 1.01 (0.57, 1.78) | 0.968 | 1.00 (0.56, 1.79) | 0.991 | 1.03 (0.58, 1.85) | 0.915 | 1.03 (0.57, 1.84) | 0.933 | 0.99 (0.55, 1.78) | 0.964 |
| Urgency | ICD-10 depression | 2.43 (1.56, 3.78) | <.001 | 2.33 (1.49, 3.65) | <.001 | 2.00 (1.25, 3.19) | 0.004 | 2.08 (1.30, 3.34) | 0.002 | 2.05 (1.28, 3.30) | 0.003 | 1.94 (1.19, 3.14) | 0.008 |
| Frequent urination | ICD-10 depression | 1.33 (0.67, 2.67) | 0.413 | 1.26 (0.63, 2.54) | 0.516 | 1.08 (0.53, 2.22) | 0.822 | 1.12 (0.55, 2.30) | 0.752 | 1.11 (0.54, 2.28) | 0.774 | 1.04 (0.50, 2.14) | 0.919 |
| Low voided volume | ICD-10 depression | 1.82 (1.10, 2.99) | 0.019 | 1.69 (1.02, 2.81) | 0.041 | 1.60 (0.95, 2.67) | 0.075 | 1.62 (0.97, 2.71) | 0.064 | 1.62 (0.97, 2.70) | 0.067 | 1.53 (0.91, 2.57) | 0.107 |
| Voiding postponement | ICD-10 depression | 1.74 (1.29, 2.35) | <.001 | 1.70 (1.25, 2.31) | 0.001 | 1.61 (1.18, 2.20) | 0.003 | 1.62 (1.18, 2.22) | 0.003 | 1.61 (1.18, 2.21) | 0.003 | 1.58 (1.15, 2.16) | 0.005 |
| Nocturia | ICD-10 depression | 1.32 (0.89, 1.96) | 0.163 | 1.29 (0.86, 1.92) | 0.212 | 1.16 (0.77, 1.75) | 0.472 | 1.19 (0.79, 1.80) | 0.398 | 1.19 (0.79, 1.79) | 0.414 | 1.11 (0.73, 1.69) | 0.628 |
| **Exposure** | **Outcome** | **OR (95% CI)** | **p** | **OR (95% CI)** | **p** | **OR (95% CI)** | **p** | **OR (95% CI)** | **p** | **OR (95% CI)** | **p** | **OR (95% CI)** | **p** |
| Daytime wetting | High depressive symptoms | 2.26 (1.54, 3.32) | <.001 | 2.03 (1.38, 2.98) | <.001 | 1.95 (1.32, 2.87) | 0.001 | 1.91 (1.29, 2.84) | 0.001 | 1.84 (1.24, 2.74) | 0.002 | 1.70 (1.13, 2.56) | 0.011 |
| Bedwetting | High depressive symptoms | 1.42 (0.88, 2.32) | 0.154 | 1.48 (0.90, 2.41) | 0.12 | 1.29 (0.77, 2.15) | 0.324 | 1.26 (0.75, 2.11) | 0.381 | 1.25 (0.74, 2.11) | 0.403 | 1.10 (0.63, 1.89) | 0.741 |
| Soiling | High depressive symptoms | 1.83 (1.30, 2.59) | 0.001 | 1.68 (1.19, 2.38) | 0.004 | 1.70 (1.19, 2.43) | 0.004 | 1.59 (1.11, 2.29) | 0.012 | 1.56 (1.08, 2.25) | 0.018 | 1.52 (1.05, 2.20) | 0.029 |
| Urgency | High depressive symptoms | 1.27 (0.86, 1.90) | 0.231 | 1.23 (0.83, 1.85) | 0.302 | 1.05 (0.69, 1.61) | 0.821 | 0.97 (0.63, 1.50) | 0.905 | 0.94 (0.61, 1.45) | 0.771 | 0.87 (0.56, 1.35) | 0.535 |
| Frequent urination | High depressive symptoms | 1.35 (0.85, 2.13) | 0.202 | 1.30 (0.82, 2.07) | 0.262 | 1.16 (0.72, 1.87) | 0.543 | 1.09 (0.67, 1.76) | 0.727 | 1.06 (0.66, 1.73) | 0.799 | 1.01 (0.62, 1.64) | 0.981 |
| Low voided volume | High depressive symptoms | 2.21 (1.55, 3.14) | <.001 | 2.13 (1.49, 3.03) | <.001 | 2.02 (1.41, 2.91) | <.001 | 1.98 (1.37, 2.86) | <.001 | 1.95 (1.34, 2.82) | <.001 | 1.87 (1.28, 2.73) | 0.001 |
| Voiding postponement | High depressive symptoms | 1.68 (1.35, 2.10) | <.001 | 1.66 (1.33, 2.07) | <.001 | 1.59 (1.26, 1.99) | <.001 | 1.58 (1.25, 1.99) | <.001 | 1.56 (1.24, 1.97) | <.001 | 1.53 (1.21, 1.93) | <.001 |
| Nocturia | High depressive symptoms | 1.55 (1.19, 2.03) | 0.001 | 1.53 (1.17, 2.01) | 0.002 | 1.41 (1.07, 1.86) | 0.015 | 1.34 (1.02, 1.78) | 0.038 | 1.33 (1.00, 1.76) | 0.047 | 1.26 (0.95, 1.68) | 0.109 |
| **Exposure** | **Outcome** | **OR (95% CI)** | **p** | **OR (95% CI)** | **p** | **OR (95% CI)** | **p** | **OR (95% CI)** | **p** | **OR (95% CI)** | **p** | **OR (95% CI)** | **p** |
| Daytime wetting | GAD symptoms | 4.11 (2.51, 6.74) | <.001 | 3.57 (2.17, 5.87) | <.001 | 3.30 (1.98, 5.49) | <.001 | 3.30 (1.97, 5.51) | <.001 | 3.18 (1.89, 5.33) | <.001 | 3.01 (1.78, 5.09) | <.001 |
| Bedwetting | GAD symptoms | 2.50 (1.22, 5.09) | 0.012 | 2.64 (1.29, 5.40) | 0.008 | 2.39 (1.16, 4.92) | 0.019 | 2.36 (1.15, 4.85) | 0.02 | 2.32 (1.12, 4.78) | 0.023 | 2.16 (1.03, 4.52) | 0.042 |
| Soiling | GAD symptoms | 1.69 (0.96, 2.96) | 0.067 | 1.49 (0.84, 2.62) | 0.168 | 1.44 (0.82, 2.53) | 0.203 | 1.39 (0.79, 2.44) | 0.253 | 1.35 (0.76, 2.39) | 0.304 | 1.32 (0.74, 2.35) | 0.348 |
| Urgency | GAD symptoms | 2.64 (1.54, 4.52) | <.001 | 2.54 (1.47, 4.39) | 0.001 | 2.31 (1.33, 4.03) | 0.003 | 2.24 (1.29, 3.90) | 0.005 | 2.16 (1.23, 3.79) | 0.008 | 2.09 (1.19, 3.67) | 0.011 |
| Frequent urination | GAD symptoms | 2.80 (1.48, 5.30) | 0.002 | 2.69 (1.42, 5.10) | 0.003 | 2.56 (1.34, 4.88) | 0.005 | 2.48 (1.30, 4.74) | 0.006 | 2.44 (1.27, 4.69) | 0.008 | 2.37 (1.23, 4.57) | 0.01 |

| | | | | | | | | | | | | | |
|---|---|---|---|---|---|---|---|---|---|---|---|---|---|
| Low voided volume | GAD symptoms | 1.22 (0.62, 2.42) | 0.564 | 1.14 (0.57, 2.28) | 0.701 | 1.11 (0.56, 2.20) | 0.767 | 1.09 (0.55, 2.18) | 0.805 | 1.07 (0.53, 2.16) | 0.841 | 1.03 (0.51, 2.07) | 0.935 |
| Voiding postponement | GAD symptoms | 1.74 (1.22, 2.50) | 0.003 | 1.70 (1.19, 2.45) | 0.004 | 1.64 (1.14, 2.36) | 0.008 | 1.63 (1.13, 2.36) | 0.009 | 1.61 (1.12, 2.33) | 0.01 | 1.59 (1.10, 2.30) | 0.013 |
| Nocturia | GAD symptoms | 2.02 (1.31, 3.09) | 0.001 | 1.98 (1.28, 3.04) | 0.002 | 1.87 (1.21, 2.90) | 0.005 | 1.83 (1.18, 2.82) | 0.007 | 1.80 (1.17, 2.78) | 0.008 | 1.73 (1.12, 2.68) | 0.014 |
| **Exposure** | **Outcome** | **OR (95% CI)** | **p** | **OR (95% CI)** | **p** | **OR (95% CI)** | **p** | **OR (95% CI)** | **p** | **OR (95% CI)** | **p** | **OR (95% CI)** | **p** |
| Daytime wetting | Self-harm act | 1.66 (0.94, 2.95) | 0.082 | 1.37 (0.77, 2.46) | 0.282 | 1.26 (0.69, 2.28) | 0.449 | 1.23 (0.68, 2.24) | 0.498 | 1.16 (0.63, 2.12) | 0.637 | 1.07 (0.58, 1.98) | 0.826 |
| Bedwetting | Self-harm act | 1.98 (1.11, 3.54) | 0.021 | 2.12 (1.17, 3.83) | 0.013 | 1.94 (1.06, 3.53) | 0.031 | 1.92 (1.05, 3.50) | 0.034 | 1.87 (1.02, 3.44) | 0.044 | 1.71 (0.93, 3.16) | 0.086 |
| Soiling | Self-harm act | 1.36 (0.83, 2.22) | 0.216 | 1.17 (0.71, 1.92) | 0.541 | 1.14 (0.69, 1.88) | 0.619 | 1.11 (0.67, 1.83) | 0.691 | 1.07 (0.64, 1.77) | 0.802 | 1.04 (0.62, 1.73) | 0.885 |
| Urgency | Self-harm act | 0.70 (0.34, 1.45) | 0.337 | 0.66 (0.31, 1.38) | 0.265 | 0.57 (0.28, 1.20) | 0.138 | 0.56 (0.27, 1.17) | 0.121 | 0.52 (0.25, 1.10) | 0.089 | 0.49 (0.23, 1.05) | 0.065 |
| Frequent urination | Self-harm act | 1.63 (0.89, 2.98) | 0.112 | 1.54 (0.83, 2.85) | 0.167 | 1.43 (0.77, 2.67) | 0.255 | 1.40 (0.75, 2.62) | 0.284 | 1.37 (0.73, 2.58) | 0.33 | 1.32 (0.70, 2.50) | 0.396 |
| Low voided volume | Self-harm act | 0.88 (0.50, 1.55) | 0.645 | 0.80 (0.45, 1.43) | 0.456 | 0.77 (0.43, 1.37) | 0.377 | 0.76 (0.42, 1.35) | 0.346 | 0.74 (0.42, 1.33) | 0.313 | 0.71 (0.39, 1.27) | 0.245 |
| Voiding postponement | Self-harm act | 1.17 (0.85, 1.63) | 0.335 | 1.14 (0.82, 1.59) | 0.437 | 1.09 (0.78, 1.53) | 0.604 | 1.10 (0.78, 1.54) | 0.593 | 1.08 (0.77, 1.52) | 0.665 | 1.06 (0.75, 1.49) | 0.752 |
| Nocturia | Self-harm act | 1.35 (0.92, 1.97) | 0.126 | 1.31 (0.89, 1.93) | 0.173 | 1.24 (0.84, 1.83) | 0.288 | 1.22 (0.82, 1.80) | 0.319 | 1.20 (0.81, 1.77) | 0.373 | 1.14 (0.77, 1.70) | 0.513 |
| **Exposure** | **Outcome** | **OR (95% CI)** | **p** | **OR (95% CI)** | **p** | **OR (95% CI)** | **p** | **OR (95% CI)** | **p** | **OR (95% CI)** | **p** | **OR (95% CI)** | **p** |
| Daytime wetting | Any disordered eating | 2.06 (1.41, 3.01) | <.001 | 1.58 (1.06, 2.35) | 0.024 | 1.56 (1.05, 2.31) | 0.029 | 1.61 (1.08, 2.41) | 0.019 | 1.58 (1.06, 2.37) | 0.026 | 1.48 (0.98, 2.25) | 0.064 |
| Bedwetting | Any disordered eating | 1.51 (0.98, 2.30) | 0.059 | 1.71 (1.09, 2.69) | 0.021 | 1.72 (1.09, 2.72) | 0.02 | 1.74 (1.10, 2.76) | 0.017 | 1.72 (1.09, 2.72) | 0.02 | 1.61 (1.00, 2.59) | 0.052 |
| Soiling | Any disordered eating | 1.68 (1.21, 2.33) | 0.002 | 1.35 (0.96, 1.92) | 0.088 | 1.34 (0.94, 1.91) | 0.101 | 1.38 (0.97, 1.98) | 0.073 | 1.36 (0.95, 1.94) | 0.091 | 1.33 (0.92, 1.91) | 0.131 |
| Urgency | Any disordered eating | 1.33 (0.92, 1.94) | 0.132 | 1.25 (0.83, 1.87) | 0.278 | 1.24 (0.82, 1.88) | 0.303 | 1.27 (0.84, 1.94) | 0.259 | 1.25 (0.82, 1.90) | 0.296 | 1.21 (0.79, 1.86) | 0.378 |
| Frequent urination | Any disordered eating | 1.36 (0.90, 2.07) | 0.144 | 1.27 (0.81, 2.00) | 0.304 | 1.27 (0.80, 2.03) | 0.305 | 1.30 (0.82, 2.08) | 0.264 | 1.29 (0.81, 2.07) | 0.282 | 1.24 (0.77, 2.00) | 0.368 |
| Low voided volume | Any disordered eating | 1.66 (1.11, 2.48) | 0.014 | 1.52 (0.98, 2.36) | 0.061 | 1.53 (0.98, 2.38) | 0.061 | 1.56 (1.00, 2.44) | 0.05 | 1.56 (1.00, 2.45) | 0.05 | 1.52 (0.97, 2.40) | 0.07 |
| Voiding postponement | Any disordered eating | 1.41 (1.13, 1.75) | 0.003 | 1.38 (1.09, 1.74) | 0.007 | 1.38 (1.09, 1.74) | 0.007 | 1.37 (1.09, 1.73) | 0.008 | 1.36 (1.08, 1.72) | 0.01 | 1.36 (1.07, 1.74) | 0.013 |
| Nocturia | Any disordered eating | 1.76 (1.34, 2.31) | <.001 | 1.76 (1.32, 2.36) | <.001 | 1.78 (1.32, 2.38) | <.001 | 1.81 (1.34, 2.43) | <.001 | 1.80 (1.33, 2.42) | <.001 | 1.72 (1.27, 2.34) | 0.001 |

GAD – Generalised Anxiety Disorder; ICD-10 - International Classification of Diseases, Tenth Revision.

Adjusted 1 confounders: sex. Adjusted 2 confounders: sex and socioeconomic indicators (parental occupational social class, maternal educational attainment, family size, ethnicity, home ownership status, material hardship). Adjusted 3 confounders: sex, socioeconomic indicators, child IQ and developmental level. Adjusted 4 confounders: sex, socioeconomic indicators, child IQ, developmental level, maternal stressful life events, maternal depression and maternal anxiety. Adjusted 5 confounders: sex, socioeconomic indicators, child IQ, developmental level, maternal stressful life events, maternal depression, maternal anxiety, child Body Mass Index and earlier emotional/behaviour problems (fully adjusted model).

**Table S7. Sequential adjustments for primary outcomes using complete case data (mental health outcomes n=1,528; disordered eating outcome n=1,375)**

| | | Unadjusted | | Adjusted 1 | | Adjusted 2 | | Adjusted 3 | | Adjusted 4 | | Adjusted 5 | |
|---|---|---|---|---|---|---|---|---|---|---|---|---|---|
| **Exposure** | **Outcome** | **OR (95% CI)** | **p** | **OR (95% CI)** | **p** | **OR (95% CI)** | **p** | **OR (95% CI)** | **p** | **OR (95% CI)** | **p** | **OR (95% CI)** | **p** |
| Daytime wetting | Common mental disorder | 2.44 (1.30, 4.56) | 0.005 | 2.08 (1.10, 3.92) | 0.024 | 2.09 (1.10, 3.95) | 0.024 | 2.07 (1.09, 3.93) | 0.025 | 2.02 (1.06, 3.84) | 0.032 | 1.95 (1.02, 3.74) | 0.044 |
| Bedwetting | Common mental disorder | 1.01 (0.39, 2.60) | 0.991 | 1.13 (0.43, 2.97) | 0.800 | 1.07 (0.41, 2.84) | 0.884 | 1.07 (0.40, 2.83) | 0.891 | 1.09 (0.41, 2.90) | 0.865 | 0.89 (0.33, 2.41) | 0.820 |
| Soiling | Common mental disorder | 1.63 (0.87, 3.04) | 0.125 | 1.38 (0.74, 2.60) | 0.314 | 1.34 (0.71, 2.53) | 0.371 | 1.32 (0.70, 2.50) | 0.393 | 1.35 (0.71, 2.56) | 0.361 | 1.31 (0.69, 2.50) | 0.415 |
| Urgency | Common mental disorder | 2.23 (1.22, 4.07) | 0.009 | 2.15 (1.17, 3.97) | 0.014 | 2.09 (1.13, 3.87) | 0.019 | 2.08 (1.13, 3.86) | 0.019 | 2.04 (1.10, 3.79) | 0.024 | 2.00 (1.07, 3.73) | 0.030 |
| Frequent urination | Common mental disorder | 1.62 (0.70, 3.73) | 0.260 | 1.54 (0.66, 3.61) | 0.315 | 1.52 (0.64, 3.57) | 0.340 | 1.50 (0.64, 3.54) | 0.351 | 1.51 (0.64, 3.57) | 0.347 | 1.44 (0.60, 3.43) | 0.412 |
| Low voided volume | Common mental disorder | 1.21 (0.61, 2.41) | 0.588 | 1.10 (0.55, 2.20) | 0.798 | 1.11 (0.55, 2.23) | 0.776 | 1.10 (0.55, 2.22) | 0.785 | 1.07 (0.53, 2.17) | 0.847 | 0.98 (0.48, 1.99) | 0.945 |
| Voiding postponement | Common mental disorder | 1.60 (1.05, 2.42) | 0.028 | 1.59 (1.04, 2.43) | 0.031 | 1.56 (1.02, 2.39) | 0.039 | 1.57 (1.03, 2.40) | 0.037 | 1.48 (0.96, 2.27) | 0.073 | 1.45 (0.94, 2.23) | 0.092 |
| Nocturia | Common mental disorder | 1.29 (0.79, 2.11) | 0.309 | 1.30 (0.79, 2.14) | 0.299 | 1.30 (0.79, 2.14) | 0.308 | 1.29 (0.78, 2.13) | 0.322 | 1.28 (0.77, 2.11) | 0.344 | 1.26 (0.75, 2.09) | 0.381 |
| **Exposure** | **Outcome** | **OR (95% CI)** | **p** | **OR (95% CI)** | **p** | **OR (95% CI)** | **p** | **OR (95% CI)** | **p** | **OR (95% CI)** | **p** | **OR (95% CI)** | **p** |
| Daytime wetting | ICD-10 depression | 3.22 (1.58, 6.60) | 0.001 | 2.85 (1.38, 5.87) | 0.004 | 2.98 (1.44, 6.16) | 0.003 | 3.03 (1.46, 6.28) | 0.003 | 3.04 (1.46, 6.33) | 0.003 | 2.94 (1.40, 6.16)† | 0.004 |
| Bedwetting | ICD-10 depression | 0.35 (0.05, 2.58) | 0.303 | 0.38 (0.05, 2.81) | 0.344 | 0.36 (0.05, 2.64) | 0.313 | 0.36 (0.05, 2.66) | 0.315 | 0.36 (0.05, 2.71) | 0.323 | 0.30 (0.04, 2.27) † | 0.245 |
| Soiling | ICD-10 depression | 1.57 (0.70, 3.52) | 0.275 | 1.38 (0.61, 3.11) | 0.438 | 1.36 (0.60, 3.08) | 0.468 | 1.39 (0.61, 3.16) | 0.437 | 1.38 (0.61, 3.15) | 0.442 | 1.35 (0.59, 3.10)† | 0.477 |
| Urgency | ICD-10 depression | 3.09 (1.56, 6.11) | 0.001 | 2.99 (1.50, 5.95) | 0.002 | 2.89 (1.44, 5.80) | 0.003 | 2.93 (1.46, 5.87) | 0.002 | 2.95 (1.47, 5.93) | 0.002 | 2.90 (1.43, 5.85)† | 0.003 |
| Frequent urination | ICD-10 depression | 1.20 (0.36, 3.98) | 0.764 | 1.15 (0.35, 3.83) | 0.818 | 1.15 (0.34, 3.85) | 0.824 | 1.17 (0.35, 3.92) | 0.802 | 1.18 (0.35, 3.95) | 0.791 | 1.13 (0.33, 3.81)† | 0.850 |
| Low voided volume | ICD-10 depression | 1.91 (0.89, 4.10) | 0.099 | 1.77 (0.82, 3.83) | 0.145 | 1.79 (0.82, 3.89) | 0.141 | 1.81 (0.83, 3.93) | 0.136 | 1.79 (0.82, 3.90) | 0.142 | 1.63 (0.74, 3.58)† | 0.221 |
| Voiding postponement | ICD-10 depression | 1.40 (0.80, 2.44) | 0.237 | 1.39 (0.79, 2.43) | 0.251 | 1.36 (0.78, 2.39) | 0.282 | 1.36 (0.77, 2.39) | 0.287 | 1.35 (0.77, 2.38) | 0.298 | 1.30 (0.74, 2.30)† | 0.362 |
| Nocturia | ICD-10 depression | 1.51 (0.82, 2.77) | 0.187 | 1.52 (0.82, 2.80) | 0.184 | 1.52 (0.82, 2.82) | 0.180 | 1.54 (0.83, 2.85) | 0.169 | 1.55 (0.83, 2.86) | 0.167 | 1.52 (0.82, 2.83)† | 0.185 |
| **Exposure** | **Outcome** | **OR (95% CI)** | **p** | **OR (95% CI)** | **p** | **OR (95% CI)** | **p** | **OR (95% CI)** | **p** | **OR (95% CI)** | **p** | **OR (95% CI)** | **p** |
| Daytime wetting | High depressive symptoms | 1.88 (1.03, 3.41) | 0.038 | 1.70 (0.93, 3.11) | 0.083 | 1.71 (0.93, 3.14) | 0.084 | 1.65 (0.89, 3.03) | 0.110 | 1.61 (0.87, 2.97) | 0.128 | 1.58 (0.86, 2.93) | 0.143 |
| Bedwetting | High depressive symptoms | 0.97 (0.43, 2.23) | 0.951 | 1.04 (0.45, 2.39) | 0.926 | 0.96 (0.41, 2.24) | 0.926 | 0.93 (0.40, 2.19) | 0.871 | 0.92 (0.39, 2.17) | 0.853 | 0.83 (0.35, 1.99) | 0.682 |
| Soiling | High depressive symptoms | 2.18 (1.29, 3.68) | 0.004 | 1.99 (1.17, 3.37) | 0.011 | 1.97 (1.15, 3.37) | 0.014 | 1.81 (1.05, 3.12) | 0.031 | 1.84 (1.07, 3.17) | 0.027 | 1.83 (1.06, 3.16) | 0.030 |
| Urgency | High depressive symptoms | 1.05 (0.55, 2.00) | 0.880 | 1.02 (0.53, 1.94) | 0.957 | 0.95 (0.49, 1.82) | 0.868 | 0.92 (0.48, 1.79) | 0.815 | 0.88 (0.45, 1.71) | 0.707 | 0.88 (0.45, 1.70) | 0.695 |
| Frequent urination | High depressive symptoms | 1.24 (0.56, 2.73) | 0.600 | 1.20 (0.54, 2.66) | 0.656 | 1.17 (0.52, 2.63) | 0.712 | 1.11 (0.49, 2.53) | 0.801 | 1.09 (0.48, 2.49) | 0.842 | 1.07 (0.46, 2.45) | 0.880 |
| Low voided volume | High depressive symptoms | 1.82 (1.05, 3.15) | 0.032 | 1.73 (0.99, 3.00) | 0.052 | 1.76 (1.01, 3.08) | 0.046 | 1.72 (0.98, 3.02) | 0.059 | 1.68 (0.96, 2.95) | 0.072 | 1.60 (0.91, 2.83) | 0.103 |
| Voiding postponement | High depressive symptoms | 1.55 (1.07, 2.24) | 0.020 | 1.54 (1.07, 2.24) | 0.022 | 1.51 (1.04, 2.21) | 0.031 | 1.55 (1.06, 2.26) | 0.023 | 1.49 (1.02, 2.19) | 0.040 | 1.47 (1.00, 2.16) | 0.047 |
| Nocturia | High depressive symptoms | 1.37 (0.90, 2.09) | 0.146 | 1.38 (0.90, 2.11) | 0.142 | 1.37 (0.89, 2.11) | 0.152 | 1.32 (0.85, 2.03) | 0.217 | 1.32 (0.85, 2.05) | 0.213 | 1.31 (0.85, 2.04) | 0.222 |
| **Exposure** | **Outcome** | **OR (95% CI)** | **p** | **OR (95% CI)** | **p** | **OR (95% CI)** | **p** | **OR (95% CI)** | **p** | **OR (95% CI)** | **p** | **OR (95% CI)** | **p** |
| Daytime wetting | GAD symptoms | 3.41 (1.55, 7.50) | 0.002 | 2.96 (1.34, 6.54) | 0.007 | 3.02 (1.36, 6.71) | 0.007 | 2.98 (1.34, 6.64) | 0.008 | 2.87 (1.28, 6.42) | 0.010 | 2.80 (1.24, 6.32)† | 0.013 |
| Bedwetting | GAD symptoms | 2.19 (0.76, 6.31) | 0.149 | 2.46 (0.84, 7.18) | 0.100 | 2.34 (0.79, 6.88) | 0.123 | 2.31 (0.79, 6.82) | 0.128 | 2.29 (0.78, 6.78) | 0.134 | 2.04 (0.68, 6.11)† | 0.203 |
| Soiling | GAD symptoms | 1.48 (0.58, 3.80) | 0.411 | 1.28 (0.50, 3.29) | 0.612 | 1.25 (0.48, 3.24) | 0.648 | 1.22 (0.47, 3.16) | 0.689 | 1.25 (0.48, 3.24) | 0.648 | 1.25 (0.48, 3.26)† | 0.649 |

| | | Unadjusted | | Adjusted 1 | | Adjusted 2 | | Adjusted 3 | | Adjusted 4 | | Adjusted 5 | |
|---|---|---|---|---|---|---|---|---|---|---|---|---|---|
| Urgency | GAD symptoms | 2.45 (1.08, 5.58) | 0.032 | 2.36 (1.03, 5.39) | 0.042 | 2.32 (1.00, 5.34) | 0.049 | 2.30 (1.00, 5.30) | 0.051 | 2.21 (0.95, 5.11) | 0.065 | 2.20 (0.95, 5.13)† | 0.066 |
| Frequent urination | GAD symptoms | 3.81 (1.54, 9.44) | 0.004 | 3.69 (1.48, 9.19) | 0.005 | 3.66 (1.45, 9.25) | 0.006 | 3.61 (1.43, 9.15) | 0.007 | 3.60 (1.41, 9.14) | 0.007 | 3.44 (1.34, 8.81)† | 0.010 |
| Low voided volume | GAD symptoms | 1.53 (0.60, 3.93) | 0.373 | 1.40 (0.55, 3.62) | 0.481 | 1.42 (0.55, 3.66) | 0.473 | 1.40 (0.54, 3.62) | 0.490 | 1.37 (0.53, 3.56) | 0.518 | 1.27 (0.49, 3.31)† | 0.629 |
| Voiding postponement | GAD symptoms | 1.12 (0.57, 2.22) | 0.741 | 1.11 (0.56, 2.20) | 0.769 | 1.08 (0.54, 2.16) | 0.823 | 1.09 (0.55, 2.18) | 0.802 | 1.03 (0.51, 2.06) | 0.934 | 1.00 (0.50, 2.00)† | 0.991 |
| Nocturia | GAD symptoms | 1.56 (0.78, 3.10) | 0.206 | 1.57 (0.79, 3.13) | 0.202 | 1.57 (0.78, 3.14) | 0.205 | 1.55 (0.77, 3.10) | 0.221 | 1.52 (0.76, 3.07) | 0.238 | 1.51 (0.75, 3.06)† | 0.246 |
| **Exposure** | **Outcome** | **OR (95% CI)** | **p** | **OR (95% CI)** | **p** | **OR (95% CI)** | **p** | **OR (95% CI)** | **p** | **OR (95% CI)** | **p** | **OR (95% CI)** | **p** |
| Daytime wetting | Self-harm act | 1.42 (0.60, 3.38) | 0.430 | 1.23 (0.51, 2.94) | 0.645 | 1.29 (0.54, 3.10) | 0.572 | 1.28 (0.53, 3.09) | 0.583 | 1.21 (0.50, 2.92) | 0.678 | 1.22 (0.50, 2.98)† | 0.657 |
| Bedwetting | Self-harm act | 2.59 (1.12, 5.99) | 0.026 | 2.91 (1.24, 6.81) | 0.014 | 2.99 (1.27, 7.08) | 0.013 | 2.99 (1.26, 7.07) | 0.013 | 2.98 (1.25, 7.12) | 0.014 | 2.77 (1.15, 6.67)† | 0.023 |
| Soiling | Self-harm act | 1.53 (0.72, 3.28) | 0.272 | 1.34 (0.62, 2.88) | 0.459 | 1.27 (0.59, 2.77) | 0.541 | 1.26 (0.58, 2.75) | 0.565 | 1.31 (0.60, 2.86) | 0.494 | 1.32 (0.61, 2.90)† | 0.482 |
| Urgency | Self-harm act | 0.56 (0.17, 1.81) | 0.334 | 0.53 (0.16, 1.73) | 0.295 | 0.51 (0.16, 1.67) | 0.266 | 0.51 (0.16, 1.67) | 0.266 | 0.48 (0.15, 1.57) | 0.224 | 0.49 (0.15, 1.61)† | 0.238 |
| Frequent urination | Self-harm act | 1.01 (0.31, 3.33) | 0.989 | 0.96 (0.29, 3.20) | 0.950 | 0.92 (0.27, 3.09) | 0.890 | 0.92 (0.27, 3.09) | 0.888 | 0.89 (0.26, 3.04) | 0.855 | 0.88 (0.26, 3.01)† | 0.833 |
| Low voided volume | Self-harm act | 0.92 (0.36, 2.33) | 0.857 | 0.84 (0.33, 2.15) | 0.720 | 0.84 (0.33, 2.15) | 0.719 | 0.83 (0.32, 2.13) | 0.699 | 0.81 (0.32, 2.09) | 0.666 | 0.76 (0.30, 1.97)† | 0.576 |
| Voiding postponement | Self-harm act | 1.15 (0.66, 1.99) | 0.624 | 1.13 (0.65, 1.97) | 0.654 | 1.09 (0.62, 1.90) | 0.769 | 1.10 (0.63, 1.92) | 0.747 | 1.01 (0.58, 1.78) | 0.963 | 0.99 (0.56, 1.74)† | 0.970 |
| Nocturia | Self-harm act | 1.24 (0.68, 2.27) | 0.481 | 1.25 (0.68, 2.29) | 0.474 | 1.23 (0.67, 2.27) | 0.504 | 1.22 (0.66, 2.26) | 0.517 | 1.19 (0.64, 2.20) | 0.588 | 1.20 (0.65, 2.23)† | 0.560 |
| **Exposure** | **Outcome** | **OR (95% CI)** | **p** | **OR (95% CI)** | **p** | **OR (95% CI)** | **p** | **OR (95% CI)** | **p** | **OR (95% CI)** | **p** | **OR (95% CI)** | **p** |
| Daytime wetting | Any disordered eating | 2.07 (1.15, 3.73) | 0.016 | 1.67 (0.91, 3.08) | 0.100 | 1.72 (0.93, 3.20) | 0.085 | 1.78 (0.96, 3.32) | 0.068 | 1.75 (0.94, 3.27) | 0.077 | 1.58 (0.84, 2.99) | 0.160 |
| Bedwetting | Any disordered eating | 1.45 (0.74, 2.85) | 0.275 | 1.66 (0.81, 3.39) | 0.163 | 1.64 (0.80, 3.37) | 0.177 | 1.68 (0.82, 3.44) | 0.158 | 1.67 (0.81, 3.44) | 0.163 | 1.41 (0.67, 2.98) | 0.365 |
| Soiling | Any disordered eating | 1.35 (0.82, 2.22) | 0.241 | 1.17 (0.69, 1.97) | 0.559 | 1.15 (0.68, 1.94) | 0.611 | 1.19 (0.70, 2.02) | 0.526 | 1.18 (0.70, 2.01) | 0.534 | 1.12 (0.66, 1.92) | 0.674 |
| Urgency | Any disordered eating | 1.24 (0.75, 2.07) | 0.405 | 1.18 (0.69, 2.02) | 0.540 | 1.21 (0.71, 2.07) | 0.484 | 1.24 (0.72, 2.13) | 0.434 | 1.22 (0.71, 2.10) | 0.472 | 1.22 (0.70, 2.11) | 0.483 |
| Frequent urination | Any disordered eating | 1.16 (0.62, 2.16) | 0.647 | 1.13 (0.59, 2.18) | 0.712 | 1.14 (0.59, 2.21) | 0.693 | 1.17 (0.60, 2.27) | 0.639 | 1.16 (0.60, 2.25) | 0.660 | 1.08 (0.55, 2.12) | 0.830 |
| Low voided volume | Any disordered eating | 1.78 (1.08, 2.94) | 0.024 | 1.69 (1.00, 2.86) | 0.051 | 1.67 (0.98, 2.83) | 0.059 | 1.74 (1.02, 2.97) | 0.043 | 1.74 (1.02, 2.97) | 0.043 | 1.65 (0.96, 2.85) | 0.071 |
| Voiding postponement | Any disordered eating | 1.35 (0.98, 1.86) | 0.065 | 1.33 (0.95, 1.87) | 0.092 | 1.34 (0.95, 1.88) | 0.091 | 1.32 (0.94, 1.86) | 0.106 | 1.29 (0.92, 1.82) | 0.145 | 1.28 (0.90, 1.81) | 0.166 |
| Nocturia | Any disordered eating | 1.51 (1.02, 2.22) | 0.037 | 1.59 (1.06, 2.39) | 0.025 | 1.59 (1.05, 2.39) | 0.027 | 1.62 (1.07, 2.44) | 0.022 | 1.61 (1.07, 2.43) | 0.023 | 1.58 (1.04, 2.41) | 0.032 |

GAD – Generalised Anxiety Disorder; ICD-10 - International Classification of Diseases, Tenth Revision. †Analyses performed on n=1494 due to 34 observations being dropped (underweight BMI predicted outcome perfectly).

Adjusted 1 confounders: sex. Adjusted 2 confounders: sex and socioeconomic indicators (parental occupational social class, maternal educational attainment, family size, ethnicity, home ownership status, material hardship). Adjusted 3 confounders: sex, socioeconomic indicators, child IQ and developmental level. Adjusted 4 confounders: sex, socioeconomic indicators, child IQ, developmental level, maternal stressful life events, maternal depression and maternal anxiety. Adjusted 5 confounders: sex, socioeconomic indicators, child IQ, developmental level, maternal stressful life events, maternal depression, maternal anxiety, child Body Mass Index and earlier emotional/behaviour problems (fully adjusted model).

**Table S8. Sequential adjustments for secondary outcomes using imputed data (n=7,332)**

| | | Unadjusted | | Adjusted 1 | | Adjusted 2 | | Adjusted 3 | | Adjusted 4 | | Adjusted 5 | |
|---|---|---|---|---|---|---|---|---|---|---|---|---|---|
| **Exposure** | **Outcome** | **B (95% CI)** | **p** | **B (95% CI)** | **p** | **B (95% CI)** | **p** | **B (95% CI)** | **p** | **B (95% CI)** | **p** | **B (95% CI)** | **p** |
| Daytime wetting | Physical anxiety | 2.15 (0.77, 3.52) | 0.002 | 1.19 (-0.16, 2.54) | 0.083 | 1.13 (-0.20, 2.45) | 0.096 | 1.10 (-0.23, 2.43) | 0.104 | 0.99 (-0.34, 2.32) | 0.146 | 0.92 (-0.42, 2.25) | 0.177 |
| Bedwetting | Physical anxiety | 0.19 (-1.51, 1.89) | 0.826 | 0.42 (-1.23, 2.08) | 0.617 | 0.32 (-1.31, 1.95) | 0.700 | 0.30 (-1.34, 1.93) | 0.720 | 0.29 (-1.35, 1.93) | 0.731 | 0.16 (-1.48, 1.81) | 0.844 |
| Soiling | Physical anxiety | 2.26 (1.04, 3.47) | <.001 | 1.51 (0.32, 2.69) | 0.013 | 1.47 (0.29, 2.65) | 0.015 | 1.42 (0.23, 2.60) | 0.019 | 1.36 (0.17, 2.54) | 0.025 | 1.34 (0.15, 2.52) | 0.027 |
| Urgency | Physical anxiety | 1.93 (0.69, 3.16) | 0.002 | 1.67 (0.44, 2.89) | 0.008 | 1.59 (0.35, 2.82) | 0.012 | 1.53 (0.29, 2.77) | 0.016 | 1.44 (0.19, 2.68) | 0.024 | 1.36 (0.12, 2.60) | 0.031 |
| Frequent urination | Physical anxiety | 1.10 (-0.51, 2.70) | 0.179 | 0.80 (-0.77, 2.38) | 0.315 | 0.76 (-0.78, 2.31) | 0.333 | 0.71 (-0.84, 2.26) | 0.369 | 0.64 (-0.91, 2.20) | 0.416 | 0.60 (-0.94, 2.15) | 0.443 |
| Low voided volume | Physical anxiety | 1.94 (0.56, 3.31) | 0.006 | 1.56 (0.20, 2.91) | 0.024 | 1.58 (0.23, 2.92) | 0.022 | 1.55 (0.21, 2.90) | 0.024 | 1.48 (0.13, 2.83) | 0.032 | 1.43 (0.08, 2.79) | 0.039 |
| Voiding postponement | Physical anxiety | 1.89 (1.17, 2.61) | <.001 | 1.75 (1.05, 2.45) | <.001 | 1.72 (1.02, 2.41) | <.001 | 1.71 (1.02, 2.41) | <.001 | 1.67 (0.98, 2.36) | <.001 | 1.64 (0.95, 2.33) | <.001 |
| Nocturia | Physical anxiety | 1.10 (0.20, 2.00) | 0.017 | 0.95 (0.07, 1.84) | 0.035 | 0.91 (0.03, 1.79) | 0.042 | 0.87 (-0.01, 1.75) | 0.054 | 0.84 (-0.05, 1.72) | 0.064 | 0.81 (-0.07, 1.69) | 0.072 |
| **Exposure** | **Outcome** | **B (95% CI)** | **p** | **B (95% CI)** | **p** | **B (95% CI)** | **p** | **B (95% CI)** | **p** | **B (95% CI)** | **p** | **B (95% CI)** | **p** |
| Daytime wetting | Mental anxiety | 1.58 (0.65, 2.51) | 0.001 | 1.41 (0.48, 2.33) | 0.003 | 1.32 (0.40, 2.25) | 0.005 | 1.31 (0.39, 2.24) | 0.006 | 1.23 (0.30, 2.17) | 0.010 | 1.08 (0.15, 2.02) | 0.023 |
| Bedwetting | Mental anxiety | 0.65 (-0.52, 1.83) | 0.275 | 0.70 (-0.48, 1.87) | 0.243 | 0.60 (-0.57, 1.78) | 0.311 | 0.60 (-0.57, 1.78) | 0.313 | 0.59 (-0.58, 1.77) | 0.322 | 0.37 (-0.80, 1.55) | 0.531 |
| Soiling | Mental anxiety | 1.43 (0.59, 2.28) | 0.001 | 1.29 (0.45, 2.14) | 0.003 | 1.27 (0.42, 2.11) | 0.003 | 1.27 (0.42, 2.12) | 0.004 | 1.22 (0.37, 2.07) | 0.005 | 1.17 (0.32, 2.02) | 0.007 |
| Urgency | Mental anxiety | 1.00 (0.16, 1.83) | 0.019 | 0.95 (0.12, 1.78) | 0.026 | 0.85 (0.02, 1.69) | 0.045 | 0.86 (0.03, 1.69) | 0.043 | 0.79 (-0.05, 1.63) | 0.064 | 0.67 (-0.16, 1.51) | 0.115 |
| Frequent urination | Mental anxiety | 0.55 (-0.54, 1.64) | 0.320 | 0.49 (-0.59, 1.58) | 0.371 | 0.44 (-0.63, 1.50) | 0.420 | 0.43 (-0.62, 1.49) | 0.420 | 0.38 (-0.68, 1.44) | 0.479 | 0.29 (-0.77, 1.34) | 0.595 |
| Low voided volume | Mental anxiety | 1.60 (0.66, 2.54) | 0.001 | 1.53 (0.58, 2.47) | 0.002 | 1.50 (0.55, 2.44) | 0.002 | 1.50 (0.55, 2.44) | 0.002 | 1.44 (0.49, 2.39) | 0.003 | 1.36 (0.41, 2.31) | 0.005 |
| Voiding postponement | Mental anxiety | 1.45 (0.99, 1.92) | <.001 | 1.43 (0.96, 1.89) | <.001 | 1.38 (0.92, 1.84) | <.001 | 1.39 (0.93, 1.85) | <.001 | 1.36 (0.90, 1.82) | <.001 | 1.31 (0.85, 1.78) | <.001 |
| Nocturia | Mental anxiety | 0.52 (-0.18, 1.22) | 0.142 | 0.49 (-0.20, 1.19) | 0.163 | 0.44 (-0.25, 1.13) | 0.208 | 0.45 (-0.25, 1.14) | 0.204 | 0.42 (-0.27, 1.11) | 0.230 | 0.34 (-0.35, 1.03) | 0.332 |
| **Exposure** | **Outcome** | **OR (95% CI)** | **p** | **OR (95% CI)** | **p** | **OR (95% CI)** | **p** | **OR (95% CI)** | **p** | **OR (95% CI)** | **p** | **OR (95% CI)** | **p** |
| Daytime wetting | Self-harm thoughts | 2.48 (1.58, 3.91) | <.001 | 2.16 (1.36, 3.42) | 0.001 | 2.09 (1.30, 3.36) | 0.002 | 2.05 (1.27, 3.30) | 0.003 | 1.96 (1.21, 3.18) | 0.006 | 1.83 (1.12, 2.98) | 0.015 |
| Bedwetting | Self-harm thoughts | 1.36 (0.70, 2.66) | 0.360 | 1.43 (0.73, 2.80) | 0.300 | 1.31 (0.66, 2.61) | 0.443 | 1.29 (0.64, 2.58) | 0.471 | 1.26 (0.63, 2.53) | 0.511 | 1.14 (0.56, 2.31) | 0.715 |
| Soiling | Self-harm thoughts | 1.70 (1.11, 2.61) | 0.015 | 1.52 (0.98, 2.35) | 0.063 | 1.50 (0.96, 2.33) | 0.074 | 1.45 (0.93, 2.25) | 0.098 | 1.41 (0.90, 2.19) | 0.130 | 1.37 (0.88, 2.14) | 0.163 |
| Urgency | Self-harm thoughts | 1.19 (0.70, 2.03) | 0.508 | 1.14 (0.67, 1.96) | 0.620 | 1.01 (0.59, 1.73) | 0.973 | 0.98 (0.57, 1.67) | 0.930 | 0.93 (0.54, 1.62) | 0.803 | 0.88 (0.51, 1.54) | 0.661 |

| | | | | | | | | | | | | | |
|---|---|---|---|---|---|---|---|---|---|---|---|---|---|
| Frequent urination | Self-harm thoughts | 1.05 (0.55, 2.01) | 0.889 | 1.00 (0.52, 1.93) | 0.994 | 0.91 (0.47, 1.78) | 0.784 | 0.88 (0.45, 1.72) | 0.712 | 0.86 (0.44, 1.69) | 0.660 | 0.82 (0.42, 1.62) | 0.565 |
| Low voided volume | Self-harm thoughts | 1.67 (1.11, 2.52) | 0.015 | 1.58 (1.04, 2.40) | 0.032 | 1.52 (1.00, 2.32) | 0.051 | 1.49 (0.98, 2.28) | 0.064 | 1.47 (0.96, 2.25) | 0.076 | 1.42 (0.92, 2.18) | 0.111 |
| Voiding postponement | Self-harm thoughts | 1.64 (1.27, 2.12) | <.001 | 1.61 (1.24, 2.08) | <.001 | 1.56 (1.20, 2.03) | 0.001 | 1.56 (1.20, 2.04) | 0.001 | 1.54 (1.18, 2.02) | 0.002 | 1.52 (1.16, 1.99) | 0.003 |
| Nocturia | Self-harm thoughts | 1.65 (1.19, 2.28) | 0.003 | 1.62 (1.16, 2.25) | 0.004 | 1.52 (1.09, 2.11) | 0.014 | 1.49 (1.07, 2.07) | 0.018 | 1.47 (1.05, 2.04) | 0.023 | 1.40 (1.00, 1.96) | 0.047 |

| **Exposure** | **Outcome** | **OR (95% CI)** | **p** | **OR (95% CI)** | **p** | **OR (95% CI)** | **p** | **OR (95% CI)** | **p** | **OR (95% CI)** | **p** | **OR (95% CI)** | **p** |
|---|---|---|---|---|---|---|---|---|---|---|---|---|---|
| Daytime wetting | Fasting | 1.77 (1.05, 2.97) | 0.031 | 1.35 (0.80, 2.27) | 0.264 | 1.30 (0.76, 2.22) | 0.336 | 1.30 (0.76, 2.21) | 0.340 | 1.26 (0.73, 2.17) | 0.399 | 1.12 (0.65, 1.94) | 0.690 |
| Bedwetting | Fasting | 1.63 (0.87, 3.03) | 0.125 | 1.82 (0.94, 3.49) | 0.074 | 1.70 (0.87, 3.31) | 0.120 | 1.68 (0.86, 3.28) | 0.129 | 1.64 (0.84, 3.22) | 0.150 | 1.43 (0.71, 2.87) | 0.320 |
| Soiling | Fasting | 1.77 (1.17, 2.67) | 0.007 | 1.43 (0.93, 2.19) | 0.104 | 1.44 (0.93, 2.23) | 0.102 | 1.40 (0.90, 2.18) | 0.135 | 1.37 (0.88, 2.14) | 0.163 | 1.32 (0.84, 2.08) | 0.225 |
| Urgency | Fasting | 1.83 (1.16, 2.88) | 0.009 | 1.73 (1.07, 2.77) | 0.024 | 1.56 (0.96, 2.55) | 0.074 | 1.52 (0.93, 2.47) | 0.094 | 1.49 (0.91, 2.43) | 0.115 | 1.41 (0.84, 2.34) | 0.189 |
| Frequent urination | Fasting | 2.84 (1.66, 4.85) | <.001 | 2.73 (1.55, 4.81) | 0.001 | 2.60 (1.44, 4.68) | 0.002 | 2.54 (1.41, 4.56) | 0.002 | 2.53 (1.41, 4.56) | 0.002 | 2.43 (1.32, 4.46) | 0.004 |
| Low voided volume | Fasting | 1.67 (1.04, 2.68) | 0.035 | 1.51 (0.92, 2.47) | 0.104 | 1.47 (0.88, 2.45) | 0.136 | 1.46 (0.87, 2.44) | 0.147 | 1.47 (0.88, 2.48) | 0.142 | 1.39 (0.82, 2.37) | 0.220 |
| Voiding postponement | Fasting | 1.80 (1.33, 2.44) | <.001 | 1.76 (1.28, 2.41) | <.001 | 1.72 (1.25, 2.38) | 0.001 | 1.71 (1.24, 2.37) | 0.001 | 1.71 (1.23, 2.37) | 0.001 | 1.69 (1.21, 2.35) | 0.002 |
| Nocturia | Fasting | 1.99 (1.39, 2.83) | <.001 | 1.96 (1.35, 2.84) | <.001 | 1.86 (1.27, 2.72) | 0.001 | 1.82 (1.25, 2.65) | 0.002 | 1.81 (1.23, 2.64) | 0.002 | 1.69 (1.15, 2.48) | 0.008 |

| **Exposure** | **Outcome** | **OR (95% CI)** | **p** | **OR (95% CI)** | **p** | **OR (95% CI)** | **p** | **OR (95% CI)** | **p** | **OR (95% CI)** | **p** | **OR (95% CI)** | **p** |
|---|---|---|---|---|---|---|---|---|---|---|---|---|---|
| Daytime wetting | Purging | 2.77 (1.59, 4.85) | <.001 | 2.08 (1.19, 3.65) | 0.011 | 1.90 (1.06, 3.40) | 0.031 | 1.99 (1.10, 3.58) | 0.022 | 1.95 (1.07, 3.53) | 0.029 | 1.88 (1.02, 3.46) | 0.042 |
| Bedwetting | Purging | 1.82 (0.86, 3.84) | 0.118 | 2.04 (0.93, 4.48) | 0.074 | 1.87 (0.84, 4.16) | 0.124 | 1.91 (0.86, 4.24) | 0.112 | 1.84 (0.82, 4.13) | 0.139 | 1.78 (0.77, 4.11) | 0.179 |
| Soiling | Purging | 2.07 (1.22, 3.49) | 0.007 | 1.63 (0.96, 2.78) | 0.071 | 1.58 (0.92, 2.73) | 0.099 | 1.63 (0.93, 2.84) | 0.085 | 1.57 (0.90, 2.75) | 0.113 | 1.55 (0.89, 2.72) | 0.122 |
| Urgency | Purging | 1.68 (0.87, 3.21) | 0.119 | 1.55 (0.80, 3.01) | 0.194 | 1.38 (0.70, 2.72) | 0.351 | 1.41 (0.71, 2.80) | 0.326 | 1.38 (0.69, 2.77) | 0.355 | 1.37 (0.68, 2.76) | 0.378 |
| Frequent urination | Purging | 1.07 (0.44, 2.60) | 0.883 | 0.97 (0.39, 2.41) | 0.941 | 0.87 (0.35, 2.17) | 0.760 | 0.88 (0.36, 2.20) | 0.792 | 0.87 (0.35, 2.18) | 0.773 | 0.86 (0.34, 2.16) | 0.739 |
| Low voided volume | Purging | 1.88 (1.05, 3.36) | 0.032 | 1.68 (0.93, 3.03) | 0.086 | 1.66 (0.91, 3.02) | 0.100 | 1.70 (0.92, 3.12) | 0.088 | 1.71 (0.93, 3.14) | 0.083 | 1.69 (0.91, 3.13) | 0.097 |
| Voiding postponement | Purging | 1.53 (0.99, 2.35) | 0.054 | 1.47 (0.95, 2.27) | 0.085 | 1.41 (0.90, 2.22) | 0.131 | 1.40 (0.89, 2.21) | 0.144 | 1.39 (0.89, 2.20) | 0.151 | 1.39 (0.88, 2.19) | 0.152 |
| Nocturia | Purging | 1.72 (1.06, 2.79) | 0.027 | 1.66 (1.01, 2.73) | 0.044 | 1.58 (0.95, 2.63) | 0.078 | 1.60 (0.96, 2.67) | 0.070 | 1.59 (0.95, 2.65) | 0.078 | 1.55 (0.92, 2.60) | 0.101 |

| **Exposure** | **Outcome** | **OR (95% CI)** | **p** | **OR (95% CI)** | **p** | **OR (95% CI)** | **p** | **OR (95% CI)** | **p** | **OR (95% CI)** | **p** | **OR (95% CI)** | **p** |
|---|---|---|---|---|---|---|---|---|---|---|---|---|---|
| Daytime wetting | Binge-eating | 3.24 (2.13, 4.92) | <.001 | 2.59 (1.67, 4.03) | <.001 | 2.48 (1.58, 3.88) | <.001 | 2.49 (1.58, 3.92) | <.001 | 2.42 (1.53, 3.83) | <.001 | 2.26 (1.42, 3.59) | 0.001 |

| | | | | | | | | | | | | | |
|---|---|---|---|---|---|---|---|---|---|---|---|---|---|
| Bedwetting | Binge-eating | 1.29 (0.73, 2.28) | 0.387 | 1.40 (0.77, 2.53) | 0.266 | 1.32 (0.72, 2.41) | 0.366 | 1.31 (0.71, 2.40) | 0.388 | 1.27 (0.69, 2.35) | 0.436 | 1.15 (0.61, 2.14) | 0.670 |
| Soiling | Binge-eating | 2.06 (1.35, 3.14) | 0.001 | 1.71 (1.11, 2.64) | 0.016 | 1.70 (1.10, 2.63) | 0.018 | 1.65 (1.06, 2.58) | 0.026 | 1.62 (1.05, 2.52) | 0.031 | 1.58 (1.02, 2.46) | 0.040 |
| Urgency | Binge-eating | 1.69 (1.05, 2.73) | 0.031 | 1.60 (0.97, 2.63) | 0.063 | 1.51 (0.91, 2.52) | 0.109 | 1.47 (0.88, 2.46) | 0.140 | 1.42 (0.85, 2.38) | 0.176 | 1.36 (0.81, 2.28) | 0.239 |
| Frequent urination | Binge-eating | 1.18 (0.66, 2.12) | 0.582 | 1.09 (0.59, 2.00) | 0.782 | 1.03 (0.55, 1.90) | 0.935 | 1.00 (0.54, 1.85) | 0.993 | 0.98 (0.53, 1.84) | 0.960 | 0.94 (0.50, 1.75) | 0.838 |
| Low voided volume | Binge-eating | 2.04 (1.31, 3.18) | 0.002 | 1.88 (1.19, 2.97) | 0.007 | 1.84 (1.17, 2.91) | 0.009 | 1.83 (1.15, 2.90) | 0.010 | 1.85 (1.17, 2.93) | 0.009 | 1.79 (1.12, 2.85) | 0.015 |
| Voiding postponement | Binge-eating | 1.43 (1.06, 1.94) | 0.021 | 1.39 (1.02, 1.90) | 0.037 | 1.36 (1.00, 1.87) | 0.051 | 1.36 (0.99, 1.86) | 0.056 | 1.35 (0.98, 1.85) | 0.063 | 1.33 (0.97, 1.83) | 0.080 |
| Nocturia | Binge-eating | 1.65 (1.18, 2.31) | 0.004 | 1.61 (1.14, 2.29) | 0.008 | 1.58 (1.11, 2.24) | 0.012 | 1.54 (1.08, 2.19) | 0.017 | 1.52 (1.07, 2.17) | 0.020 | 1.45 (1.01, 2.07) | 0.042 |

| **Exposure** | **Outcome** | **OR (95% CI)** | **p** | **OR (95% CI)** | **p** | **OR (95% CI)** | **p** | **OR (95% CI)** | **p** | **OR (95% CI)** | **p** | **OR (95% CI)** | **p** |
|---|---|---|---|---|---|---|---|---|---|---|---|---|---|
| Daytime wetting | Excessive exercise | 1.77 (1.14, 2.76) | 0.012 | 1.34 (0.84, 2.14) | 0.214 | 1.39 (0.86, 2.24) | 0.180 | 1.46 (0.90, 2.37) | 0.123 | 1.46 (0.90, 2.36) | 0.124 | 1.40 (0.86, 2.27) | 0.179 |
| Bedwetting | Excessive exercise | 1.43 (0.88, 2.32) | 0.148 | 1.61 (0.95, 2.71) | 0.075 | 1.76 (1.03, 3.00) | 0.040 | 1.81 (1.06, 3.09) | 0.031 | 1.80 (1.05, 3.09) | 0.032 | 1.76 (1.01, 3.05) | 0.045 |
| Soiling | Excessive exercise | 1.37 (0.95, 1.97) | 0.090 | 1.09 (0.75, 1.58) | 0.659 | 1.08 (0.74, 1.57) | 0.692 | 1.15 (0.78, 1.68) | 0.484 | 1.14 (0.77, 1.67) | 0.511 | 1.11 (0.76, 1.64) | 0.583 |
| Urgency | Excessive exercise | 0.87 (0.57, 1.33) | 0.528 | 0.79 (0.51, 1.23) | 0.300 | 0.84 (0.54, 1.32) | 0.448 | 0.89 (0.56, 1.40) | 0.603 | 0.88 (0.56, 1.39) | 0.590 | 0.87 (0.55, 1.37) | 0.549 |
| Frequent urination | Excessive exercise | 1.21 (0.74, 1.99) | 0.442 | 1.11 (0.66, 1.87) | 0.687 | 1.18 (0.70, 2.01) | 0.531 | 1.25 (0.72, 2.15) | 0.425 | 1.24 (0.72, 2.14) | 0.436 | 1.21 (0.70, 2.11) | 0.491 |
| Low voided volume | Excessive exercise | 1.63 (1.08, 2.45) | 0.020 | 1.48 (0.95, 2.31) | 0.080 | 1.53 (0.98, 2.39) | 0.062 | 1.59 (1.01, 2.49) | 0.044 | 1.58 (1.00, 2.49) | 0.048 | 1.56 (0.98, 2.46) | 0.059 |
| Voiding postponement | Excessive exercise | 1.35 (1.05, 1.75) | 0.019 | 1.32 (1.01, 1.72) | 0.045 | 1.35 (1.03, 1.78) | 0.030 | 1.35 (1.02, 1.78) | 0.035 | 1.34 (1.02, 1.78) | 0.037 | 1.36 (1.03, 1.80) | 0.033 |
| Nocturia | Excessive exercise | 1.56 (1.16, 2.11) | 0.004 | 1.54 (1.12, 2.11) | 0.008 | 1.62 (1.17, 2.23) | 0.004 | 1.69 (1.22, 2.33) | 0.002 | 1.69 (1.22, 2.33) | 0.002 | 1.64 (1.18, 2.28) | 0.003 |

| **Exposure** | **Outcome** | **OR (95% CI)** | **p** | **OR (95% CI)** | **p** | **OR (95% CI)** | **p** | **OR (95% CI)** | **p** | **OR (95% CI)** | **p** | **OR (95% CI)** | **p** |
|---|---|---|---|---|---|---|---|---|---|---|---|---|---|
| Daytime wetting | DSM-5 DE | 2.35 (1.42, 3.89) | 0.001 | 1.88 (1.12, 3.14) | 0.016 | 1.99 (1.17, 3.39) | 0.012 | 2.06 (1.20, 3.54) | 0.009 | 2.03 (1.18, 3.49) | 0.011 | 1.83 (1.04, 3.20) | 0.036 |
| Bedwetting | DSM-5 DE | 1.39 (0.72, 2.70) | 0.325 | 1.51 (0.76, 3.00) | 0.235 | 1.47 (0.73, 2.98) | 0.281 | 1.47 (0.72, 2.97) | 0.286 | 1.44 (0.71, 2.92) | 0.310 | 1.25 (0.60, 2.61) | 0.546 |
| Soiling | DSM-5 DE | 2.13 (1.37, 3.32) | 0.001 | 1.79 (1.13, 2.82) | 0.013 | 1.85 (1.16, 2.94) | 0.010 | 1.81 (1.13, 2.91) | 0.013 | 1.78 (1.10, 2.86) | 0.018 | 1.72 (1.07, 2.78) | 0.026 |
| Urgency | DSM-5 DE | 1.44 (0.87, 2.37) | 0.151 | 1.35 (0.81, 2.26) | 0.244 | 1.21 (0.71, 2.04) | 0.478 | 1.17 (0.69, 1.98) | 0.559 | 1.15 (0.68, 1.95) | 0.598 | 1.09 (0.64, 1.88) | 0.742 |
| Frequent urination | DSM-5 DE | 2.26 (1.27, 4.01) | 0.006 | 2.15 (1.18, 3.89) | 0.012 | 2.01 (1.08, 3.71) | 0.027 | 1.97 (1.06, 3.64) | 0.031 | 1.94 (1.05, 3.60) | 0.034 | 1.85 (0.99, 3.47) | 0.055 |
| Low voided volume | DSM-5 DE | 1.85 (1.11, 3.08) | 0.019 | 1.70 (1.00, 2.89) | 0.049 | 1.66 (0.97, 2.85) | 0.064 | 1.68 (0.98, 2.88) | 0.061 | 1.66 (0.97, 2.86) | 0.065 | 1.59 (0.91, 2.77) | 0.099 |
| Voiding postponement | DSM-5 DE | 1.52 (1.08, 2.14) | 0.015 | 1.48 (1.05, 2.09) | 0.026 | 1.45 (1.02, 2.06) | 0.039 | 1.43 (1.01, 2.03) | 0.047 | 1.42 (1.00, 2.02) | 0.051 | 1.40 (0.98, 2.01) | 0.064 |

| | | | | | | | | | | | | | |
|---|---|---|---|---|---|---|---|---|---|---|---|---|---|
| Nocturia | DSM-5 DE | 1.56 (1.06, 2.32) | 0.026 | 1.53 (1.02, 2.29) | 0.041 | 1.43 (0.94, 2.16) | 0.091 | 1.39 (0.92, 2.11) | 0.121 | 1.38 (0.91, 2.09) | 0.131 | 1.28 (0.83, 1.97) | 0.256 |

DSM-5 DE- Disordered eating at Diagnostic and Statistical Manual of Mental Disorders, Fifth Edition frequency (at least once a week).

Adjusted 1 confounders: sex. Adjusted 2 confounders: sex and socioeconomic indicators (parental occupational social class, maternal educational attainment, family size, ethnicity, home ownership status, material hardship). Adjusted 3 confounders: sex, socioeconomic indicators, child IQ and developmental level. Adjusted 4 confounders: sex, socioeconomic indicators, child IQ, developmental level, maternal stressful life events, maternal depression and maternal anxiety. Adjusted 5 confounders: sex, socioeconomic indicators, child IQ, developmental level, maternal stressful life events, maternal depression, maternal anxiety, child Body Mass Index and earlier emotional/behaviour problems (fully adjusted model).

**Table S9. Sequential adjustments for secondary outcomes using complete case data (mental health outcomes n=1,528; disordered eating outcomes n=1,375)**

| | | Unadjusted | | Adjusted 1 | | Adjusted 2 | | Adjusted 3 | | Adjusted 4 | | Adjusted 5 | |
|---|---|---|---|---|---|---|---|---|---|---|---|---|---|
| **Exposure** | **Outcome** | **B (95% CI)** | **p** | **B (95% CI)** | **p** | **B (95% CI)** | **p** | **B (95% CI)** | **p** | **B (95% CI)** | **p** | **B (95% CI)** | **p** |
| Daytime wetting | Physical anxiety | 1.87 (-0.16, 3.89) | 0.071 | 1.18 (-0.80, 3.17) | 0.242 | 1.14 (-0.85, 3.12) | 0.261 | 1.09 (-0.89, 3.08) | 0.280 | 1.07 (-0.91, 3.06) | 0.289 | 1.12 (-0.87, 3.10) | 0.270 |
| Bedwetting | Physical anxiety | -1.28 (-3.67, 1.11) | 0.294 | -0.88 (-3.21, 1.46) | 0.463 | -1.15 (-3.49, 1.19) | 0.335 | -1.17 (-3.51, 1.18) | 0.329 | -1.13 (-3.48, 1.21) | 0.343 | -1.23 (-3.58, 1.11) | 0.304 |
| Soiling | Physical anxiety | 1.13 (-0.69, 2.94) | 0.223 | 0.47 (-1.31, 2.25) | 0.605 | 0.45 (-1.34, 2.23) | 0.624 | 0.35 (-1.44, 2.14) | 0.700 | 0.35 (-1.44, 2.14) | 0.704 | 0.35 (-1.44, 2.13) | 0.705 |
| Urgency | Physical anxiety | 2.50 (0.60, 4.39) | 0.010 | 2.29 (0.44, 4.14) | 0.015 | 2.11 (0.25, 3.96) | 0.026 | 2.09 (0.24, 3.95) | 0.027 | 2.08 (0.22, 3.94) | 0.029 | 2.11 (0.25, 3.97) | 0.026 |
| Frequent urination | Physical anxiety | 0.31 (-2.14, 2.76) | 0.805 | 0.10 (-2.30, 2.50) | 0.935 | 0.04 (-2.36, 2.44) | 0.975 | 0.00 (-2.40, 2.40) | 0.999 | -0.02 (-2.43, 2.38) | 0.984 | 0.01 (-2.39, 2.41) | 0.992 |
| Low voided volume | Physical anxiety | 1.38 (-0.47, 3.22) | 0.143 | 0.99 (-0.81, 2.80) | 0.279 | 1.10 (-0.70, 2.90) | 0.233 | 1.05 (-0.75, 2.86) | 0.252 | 1.01 (-0.80, 2.81) | 0.275 | 0.93 (-0.87, 2.74) | 0.312 |
| Voiding postponement | Physical anxiety | 1.65 (0.48, 2.82) | 0.006 | 1.60 (0.45, 2.74) | 0.006 | 1.56 (0.42, 2.71) | 0.008 | 1.59 (0.44, 2.74) | 0.007 | 1.56 (0.41, 2.71) | 0.008 | 1.52 (0.37, 2.67) | 0.010 |
| Nocturia | Physical anxiety | 1.04 (-0.29, 2.37) | 0.125 | 1.05 (-0.25, 2.35) | 0.113 | 1.06 (-0.24, 2.36) | 0.109 | 1.02 (-0.28, 2.32) | 0.124 | 1.03 (-0.27, 2.33) | 0.121 | 1.03 (-0.27, 2.33) | 0.120 |
| **Exposure** | **Outcome** | **B (95% CI)** | **p** | **B (95% CI)** | **p** | **B (95% CI)** | **p** | **B (95% CI)** | **p** | **B (95% CI)** | **p** | **B (95% CI)** | **p** |
| Daytime wetting | Mental anxiety | 0.97 (-0.32, 2.27) | 0.141 | 0.93 (-0.37, 2.23) | 0.159 | 0.87 (-0.43, 2.17) | 0.188 | 0.87 (-0.43, 2.17) | 0.190 | 0.84 (-0.46, 2.14) | 0.205 | 0.80 (-0.50, 2.10) | 0.228 |
| Bedwetting | Mental anxiety | -0.70 (-2.23, 0.83) | 0.370 | -0.67 (-2.21, 0.86) | 0.388 | -0.80 (-2.33, 0.74) | 0.308 | -0.79 (-2.32, 0.75) | 0.315 | -0.77 (-2.30, 0.76) | 0.325 | -0.94 (-2.47, 0.59) | 0.230 |
| Soiling | Mental anxiety | 1.09 (-0.07, 2.25) | 0.065 | 1.06 (-0.11, 2.22) | 0.075 | 1.03 (-0.13, 2.20) | 0.082 | 1.03 (-0.14, 2.20) | 0.085 | 1.03 (-0.14, 2.20) | 0.085 | 0.96 (-0.20, 2.13) | 0.106 |
| Urgency | Mental anxiety | 1.39 (0.18, 2.60) | 0.024 | 1.38 (0.17, 2.59) | 0.026 | 1.36 (0.15, 2.58) | 0.028 | 1.36 (0.15, 2.58) | 0.028 | 1.33 (0.12, 2.55) | 0.032 | 1.29 (0.08, 2.51) | 0.037 |
| Frequent urination | Mental anxiety | 0.33 (-1.24, 1.90) | 0.683 | 0.31 (-1.26, 1.88) | 0.696 | 0.35 (-1.22, 1.92) | 0.665 | 0.35 (-1.22, 1.93) | 0.658 | 0.33 (-1.24, 1.90) | 0.682 | 0.29 (-1.28, 1.85) | 0.722 |
| Low voided volume | Mental anxiety | 1.29 (0.11, 2.46) | 0.032 | 1.26 (0.09, 2.44) | 0.036 | 1.25 (0.07, 2.43) | 0.038 | 1.23 (0.05, 2.41) | 0.041 | 1.19 (0.01, 2.37) | 0.048 | 1.11 (-0.07, 2.29) | 0.066 |
| Voiding postponement | Mental anxiety | 1.12 (0.37, 1.87) | 0.003 | 1.12 (0.37, 1.87) | 0.003 | 1.08 (0.33, 1.84) | 0.005 | 1.09 (0.34, 1.85) | 0.004 | 1.05 (0.30, 1.81) | 0.006 | 1.02 (0.27, 1.78) | 0.008 |
| Nocturia | Mental anxiety | 0.46 (-0.39, 1.31) | 0.294 | 0.46 (-0.39, 1.31) | 0.293 | 0.45 (-0.40, 1.30) | 0.300 | 0.45 (-0.40, 1.30) | 0.300 | 0.45 (-0.40, 1.30) | 0.298 | 0.42 (-0.43, 1.27) | 0.328 |
| **Exposure** | **Outcome** | **OR (95% CI)** | **p** | **OR (95% CI)** | **p** | **OR (95% CI)** | **p** | **OR (95% CI)** | **p** | **OR (95% CI)** | **p** | **OR (95% CI)** | **p** |
| Daytime wetting | Self-harm thoughts | 1.94 (0.93, 4.05) | 0.078 | 1.69 (0.80, 3.54) | 0.168 | 1.74 (0.82, 3.68) | 0.147 | 1.72 (0.81, 3.63) | 0.159 | 1.69 (0.80, 3.58) | 0.171 | 1.63 (0.76, 3.49)† | 0.208 |
| Bedwetting | Self-harm thoughts | 1.43 (0.55, 3.71) | 0.464 | 1.58 (0.60, 4.15) | 0.350 | 1.59 (0.60, 4.22) | 0.352 | 1.58 (0.59, 4.21) | 0.361 | 1.60 (0.60, 4.27) | 0.348 | 1.39 (0.51, 3.74)† | 0.520 |
| Soiling | Self-harm thoughts | 2.35 (1.25, 4.42) | 0.008 | 2.06 (1.09, 3.90) | 0.025 | 2.02 (1.06, 3.85) | 0.032 | 1.98 (1.04, 3.78) | 0.039 | 2.00 (1.05, 3.83) | 0.035 | 1.98 (1.03, 3.81)† | 0.040 |
| Urgency | Self-harm thoughts | 1.21 (0.54, 2.71) | 0.640 | 1.16 (0.52, 2.61) | 0.720 | 1.11 (0.49, 2.51) | 0.803 | 1.11 (0.49, 2.51) | 0.807 | 1.08 (0.47, 2.46) | 0.855 | 1.05 (0.46, 2.40)† | 0.908 |

| | | Unadjusted | | Adjusted 1 | | Adjusted 2 | | Adjusted 3 | | Adjusted 4 | | Adjusted 5 | |
|---|---|---|---|---|---|---|---|---|---|---|---|---|---|
| Frequent urination | Self-harm thoughts | 1.52 (0.58, 3.96) | 0.392 | 1.46 (0.55, 3.83) | 0.445 | 1.42 (0.53, 3.79) | 0.489 | 1.40 (0.52, 3.76) | 0.502 | 1.42 (0.53, 3.80) | 0.490 | 1.36 (0.50, 3.68)† | 0.549 |
| Low voided volume | Self-harm thoughts | 1.52 (0.74, 3.14) | 0.254 | 1.41 (0.68, 2.92) | 0.359 | 1.42 (0.68, 2.97) | 0.348 | 1.39 (0.66, 2.90) | 0.385 | 1.37 (0.65, 2.87) | 0.404 | 1.26 (0.60, 2.65)† | 0.549 |
| Voiding postponement | Self-harm thoughts | 1.42 (0.88, 2.30) | 0.156 | 1.41 (0.87, 2.29) | 0.168 | 1.38 (0.84, 2.25) | 0.204 | 1.39 (0.85, 2.28) | 0.186 | 1.35 (0.82, 2.22) | 0.231 | 1.30 (0.79, 2.15)† | 0.296 |
| Nocturia | Self-harm thoughts | 1.90 (1.15, 3.14) | 0.012 | 1.92 (1.16, 3.19) | 0.011 | 1.94 (1.16, 3.23) | 0.011 | 1.92 (1.15, 3.20) | 0.013 | 1.90 (1.14, 3.18) | 0.014 | 1.90 (1.13, 3.19)† | 0.015 |
| **Exposure** | **Outcome** | **OR (95% CI)** | **p** | **OR (95% CI)** | **p** | **OR (95% CI)** | **p** | **OR (95% CI)** | **p** | **OR (95% CI)** | **p** | **OR (95% CI)** | **p** |
| Daytime wetting | Fasting | 1.01 (0.39, 2.60) | 0.981 | 0.80 (0.31, 2.08) | 0.645 | 0.85 (0.33, 2.22) | 0.739 | 0.78 (0.30, 2.06) | 0.621 | 0.74 (0.28, 1.97) | 0.552 | 0.64 (0.24, 1.71)♦ | 0.370 |
| Bedwetting | Fasting | 1.35 (0.52, 3.52) | 0.541 | 1.49 (0.55, 4.00) | 0.430 | 1.50 (0.55, 4.06) | 0.427 | 1.40 (0.51, 3.84) | 0.515 | 1.42 (0.51, 3.91) | 0.502 | 1.10 (0.38, 3.13)♦ | 0.864 |
| Soiling | Fasting | 1.85 (0.96, 3.53) | 0.064 | 1.62 (0.84, 3.14) | 0.153 | 1.73 (0.88, 3.38) | 0.111 | 1.57 (0.80, 3.09) | 0.189 | 1.57 (0.80, 3.10) | 0.193 | 1.58 (0.79, 3.13)♦ | 0.194 |
| Urgency | Fasting | 2.13 (1.13, 4.01) | 0.019 | 2.07 (1.08, 3.97) | 0.028 | 2.09 (1.08, 4.05) | 0.028 | 1.92 (0.98, 3.74) | 0.056 | 1.85 (0.94, 3.63) | 0.075 | 1.87 (0.94, 3.72)♦ | 0.073 |
| Frequent urination | Fasting | 1.60 (0.70, 3.65) | 0.267 | 1.58 (0.68, 3.68) | 0.293 | 1.54 (0.65, 3.63) | 0.322 | 1.41 (0.59, 3.36) | 0.439 | 1.40 (0.58, 3.35) | 0.453 | 1.30 (0.54, 3.13)♦ | 0.564 |
| Low voided volume | Fasting | 1.74 (0.89, 3.42) | 0.105 | 1.62 (0.81, 3.22) | 0.170 | 1.65 (0.82, 3.30) | 0.158 | 1.54 (0.76, 3.09) | 0.230 | 1.56 (0.77, 3.17) | 0.216 | 1.48 (0.72, 3.01)♦ | 0.284 |
| Voiding postponement | Fasting | 1.44 (0.91, 2.27) | 0.115 | 1.41 (0.88, 2.24) | 0.149 | 1.48 (0.92, 2.36) | 0.105 | 1.48 (0.92, 2.39) | 0.102 | 1.43 (0.89, 2.32) | 0.141 | 1.39 (0.86, 2.26)♦ | 0.178 |
| Nocturia | Fasting | 1.58 (0.92, 2.69) | 0.094 | 1.63 (0.94, 2.82) | 0.082 | 1.57 (0.90, 2.74) | 0.113 | 1.50 (0.85, 2.62) | 0.160 | 1.47 (0.84, 2.58) | 0.179 | 1.40 (0.79, 2.49)♦ | 0.245 |
| **Exposure** | **Outcome** | **OR (95% CI)** | **p** | **OR (95% CI)** | **p** | **OR (95% CI)** | **p** | **OR (95% CI)** | **p** | **OR (95% CI)** | **p** | **OR (95% CI)** | **p** |
| Daytime wetting | Purging | 2.16 (0.83, 5.64) | 0.115 | 1.74 (0.66, 4.59) | 0.261 | 1.79 (0.67, 4.76) | 0.243 | 1.81 (0.68, 4.83) | 0.236 | 1.73 (0.64, 4.64) | 0.278 | 1.58 (0.58, 4.30)♦ | 0.372 |
| Bedwetting | Purging | 1.58 (0.47, 5.26) | 0.459 | 1.73 (0.51, 5.90) | 0.380 | 1.60 (0.46, 5.53) | 0.460 | 1.65 (0.48, 5.72) | 0.430 | 1.52 (0.44, 5.32) | 0.510 | 1.30 (0.36, 4.65)♦ | 0.686 |
| Soiling | Purging | 2.06 (0.91, 4.67) | 0.084 | 1.81 (0.79, 4.14) | 0.162 | 1.86 (0.80, 4.31) | 0.148 | 1.87 (0.80, 4.36) | 0.148 | 1.91 (0.82, 4.47) | 0.134 | 1.89 (0.80, 4.45)♦ | 0.144 |
| Urgency | Purging | 1.77 (0.74, 4.24) | 0.200 | 1.69 (0.70, 4.08) | 0.247 | 1.62 (0.66, 3.96) | 0.290 | 1.60 (0.65, 3.96) | 0.305 | 1.55 (0.63, 3.85) | 0.341 | 1.55 (0.62, 3.88)♦ | 0.344 |
| Frequent urination | Purging | 1.26 (0.38, 4.17) | 0.704 | 1.23 (0.37, 4.11) | 0.740 | 1.14 (0.33, 3.86) | 0.837 | 1.14 (0.33, 3.88) | 0.835 | 1.10 (0.32, 3.80) | 0.880 | 1.01 (0.29, 3.53)♦ | 0.987 |
| Low voided volume | Purging | 2.17 (0.96, 4.93) | 0.064 | 2.01 (0.88, 4.63) | 0.099 | 2.08 (0.90, 4.82) | 0.086 | 2.10 (0.91, 4.89) | 0.084 | 2.11 (0.90, 4.92) | 0.085 | 1.98 (0.84, 4.64)♦ | 0.117 |
| Voiding postponement | Purging | 1.36 (0.73, 2.53) | 0.328 | 1.32 (0.71, 2.47) | 0.384 | 1.34 (0.72, 2.52) | 0.358 | 1.33 (0.71, 2.50) | 0.377 | 1.25 (0.66, 2.37) | 0.493 | 1.22 (0.64, 2.31)♦ | 0.546 |
| Nocturia | Purging | 2.13 (1.12, 4.08) | 0.022 | 2.20 (1.14, 4.25) | 0.019 | 2.11 (1.08, 4.11) | 0.029 | 2.11 (1.08, 4.13) | 0.029 | 2.08 (1.06, 4.08) | 0.032 | 2.01 (1.02, 3.97)♦ | 0.045 |
| **Exposure** | **Outcome** | **OR (95% CI)** | **p** | **OR (95% CI)** | **p** | **OR (95% CI)** | **p** | **OR (95% CI)** | **p** | **OR (95% CI)** | **p** | **OR (95% CI)** | **p** |
| Daytime wetting | Binge-eating | 3.14 (1.62, 6.11) | 0.001 | 2.66 (1.35, 5.23) | 0.005 | 2.74 (1.38, 5.43) | 0.004 | 2.79 (1.40, 5.54) | 0.003 | 2.71 (1.36, 5.41) | 0.005 | 2.39 (1.18, 4.83) | 0.015 |

| | | **Unadjusted** | | **Adjusted 1** | | **Adjusted 2** | | **Adjusted 3** | | **Adjusted 4** | | **Adjusted 5** | |
|---|---|---|---|---|---|---|---|---|---|---|---|---|---|
| Bedwetting | Binge-eating | 0.94 (0.33, 2.70) | 0.910 | 1.00 (0.35, 2.92) | 0.993 | 0.96 (0.33, 2.81) | 0.943 | 0.98 (0.33, 2.86) | 0.969 | 0.98 (0.33, 2.86) | 0.964 | 0.75 (0.25, 2.25) | 0.606 |
| Soiling | Binge-eating | 1.16 (0.56, 2.38) | 0.688 | 1.03 (0.49, 2.13) | 0.943 | 1.02 (0.49, 2.12) | 0.968 | 1.02 (0.49, 2.14) | 0.962 | 1.02 (0.48, 2.13) | 0.967 | 0.96 (0.45, 2.02) | 0.907 |
| Urgency | Binge-eating | 1.20 (0.58, 2.48) | 0.618 | 1.15 (0.55, 2.39) | 0.715 | 1.15 (0.55, 2.41) | 0.702 | 1.14 (0.54, 2.40) | 0.723 | 1.11 (0.52, 2.34) | 0.788 | 1.07 (0.50, 2.27) | 0.862 |
| Frequent urination | Binge-eating | 1.20 (0.50, 2.88) | 0.687 | 1.17 (0.48, 2.85) | 0.727 | 1.20 (0.49, 2.93) | 0.692 | 1.20 (0.49, 2.95) | 0.687 | 1.19 (0.48, 2.92) | 0.708 | 1.12 (0.45, 2.77) | 0.806 |
| Low voided volume | Binge-eating | 2.39 (1.31, 4.37) | 0.005 | 2.27 (1.23, 4.20) | 0.009 | 2.27 (1.22, 4.21) | 0.009 | 2.31 (1.24, 4.29) | 0.008 | 2.35 (1.26, 4.40) | 0.007 | 2.27 (1.21, 4.28) | 0.011 |
| Voiding postponement | Binge-eating | 1.29 (0.82, 2.02) | 0.273 | 1.26 (0.79, 1.99) | 0.329 | 1.27 (0.80, 2.02) | 0.307 | 1.26 (0.79, 2.00) | 0.329 | 1.21 (0.76, 1.93) | 0.418 | 1.20 (0.75, 1.91) | 0.457 |
| Nocturia | Binge-eating | 1.52 (0.90, 2.57) | 0.113 | 1.56 (0.92, 2.65) | 0.102 | 1.57 (0.92, 2.68) | 0.098 | 1.57 (0.92, 2.69) | 0.097 | 1.56 (0.91, 2.67) | 0.104 | 1.49 (0.86, 2.56) | 0.154 |
| **Exposure** | **Outcome** | **OR (95% CI)** | **p** | **OR (95% CI)** | **p** | **OR (95% CI)** | **p** | **OR (95% CI)** | **p** | **OR (95% CI)** | **p** | **OR (95% CI)** | **p** |
| Daytime wetting | Excessive exercise | 2.07 (1.12, 3.82) | 0.020 | 1.69 (0.90, 3.18) | 0.101 | 1.69 (0.89, 3.19) | 0.106 | 1.81 (0.95, 3.43) | 0.069 | 1.81 (0.96, 3.44) | 0.068 | 1.74 (0.91, 3.34) | 0.096 |
| Bedwetting | Excessive exercise | 1.15 (0.53, 2.47) | 0.721 | 1.26 (0.57, 2.79) | 0.568 | 1.27 (0.57, 2.83) | 0.561 | 1.32 (0.59, 2.96) | 0.494 | 1.31 (0.59, 2.95) | 0.506 | 1.21 (0.53, 2.78) | 0.647 |
| Soiling | Excessive exercise | 1.25 (0.72, 2.18) | 0.428 | 1.09 (0.62, 1.93) | 0.768 | 1.05 (0.59, 1.87) | 0.863 | 1.13 (0.63, 2.02) | 0.677 | 1.13 (0.63, 2.03) | 0.674 | 1.09 (0.60, 1.96) | 0.779 |
| Urgency | Excessive exercise | 0.75 (0.40, 1.43) | 0.386 | 0.70 (0.36, 1.34) | 0.280 | 0.72 (0.37, 1.39) | 0.328 | 0.76 (0.39, 1.48) | 0.427 | 0.77 (0.40, 1.49) | 0.431 | 0.77 (0.39, 1.51) | 0.448 |
| Frequent urination | Excessive exercise | 1.15 (0.57, 2.30) | 0.692 | 1.12 (0.55, 2.30) | 0.751 | 1.15 (0.56, 2.36) | 0.712 | 1.21 (0.58, 2.50) | 0.612 | 1.21 (0.58, 2.50) | 0.610 | 1.15 (0.55, 2.40) | 0.712 |
| Low voided volume | Excessive exercise | 1.56 (0.91, 2.69) | 0.107 | 1.46 (0.83, 2.56) | 0.185 | 1.44 (0.82, 2.53) | 0.206 | 1.54 (0.87, 2.72) | 0.141 | 1.52 (0.86, 2.70) | 0.152 | 1.45 (0.81, 2.58) | 0.211 |
| Voiding postponement | Excessive exercise | 1.42 (1.00, 2.02) | 0.047 | 1.41 (0.98, 2.02) | 0.065 | 1.40 (0.97, 2.01) | 0.072 | 1.38 (0.96, 1.99) | 0.084 | 1.38 (0.95, 1.99) | 0.089 | 1.37 (0.94, 1.99) | 0.098 |
| Nocturia | Excessive exercise | 1.39 (0.91, 2.13) | 0.127 | 1.44 (0.93, 2.24) | 0.105 | 1.48 (0.95, 2.31) | 0.082 | 1.55 (0.99, 2.42) | 0.055 | 1.55 (0.99, 2.43) | 0.053 | 1.55 (0.99, 2.44) | 0.057 |
| **Exposure** | **Outcome** | **OR (95% CI)** | **p** | **OR (95% CI)** | **p** | **OR (95% CI)** | **p** | **OR (95% CI)** | **p** | **OR (95% CI)** | **p** | **OR (95% CI)** | **p** |
| Daytime wetting | DSM-5 DE | 2.03 (0.93, 4.44) | 0.077 | 1.66 (0.75, 3.69) | 0.209 | 1.83 (0.82, 4.12) | 0.142 | 1.82 (0.81, 4.10) | 0.150 | 1.80 (0.80, 4.06) | 0.159 | 1.50 (0.65, 3.47) | 0.344 |
| Bedwetting | DSM-5 DE | 0.85 (0.26, 2.79) | 0.783 | 0.91 (0.27, 3.04) | 0.873 | 0.87 (0.26, 2.98) | 0.830 | 0.88 (0.26, 3.00) | 0.835 | 0.88 (0.26, 3.02) | 0.842 | 0.64 (0.18, 2.28) | 0.495 |
| Soiling | DSM-5 DE | 1.87 (0.96, 3.67) | 0.067 | 1.66 (0.84, 3.29) | 0.144 | 1.74 (0.86, 3.51) | 0.121 | 1.69 (0.83, 3.42) | 0.147 | 1.68 (0.83, 3.42) | 0.149 | 1.58 (0.77, 3.23) | 0.213 |
| Urgency | DSM-5 DE | 1.11 (0.50, 2.49) | 0.793 | 1.06 (0.47, 2.38) | 0.897 | 1.04 (0.45, 2.39) | 0.927 | 0.98 (0.42, 2.26) | 0.956 | 0.96 (0.41, 2.24) | 0.927 | 0.93 (0.40, 2.18) | 0.873 |
| Frequent urination | DSM-5 DE | 2.14 (0.98, 4.71) | 0.058 | 2.14 (0.95, 4.79) | 0.065 | 2.23 (0.98, 5.08) | 0.055 | 2.19 (0.96, 5.02) | 0.063 | 2.16 (0.94, 4.97) | 0.069 | 2.09 (0.91, 4.83) | 0.083 |
| Low voided volume | DSM-5 DE | 1.75 (0.87, 3.53) | 0.115 | 1.64 (0.80, 3.33) | 0.175 | 1.68 (0.81, 3.45) | 0.161 | 1.67 (0.81, 3.45) | 0.168 | 1.65 (0.80, 3.41) | 0.179 | 1.53 (0.73, 3.24) | 0.260 |

| | | Unadjusted | | Adjusted 1 | | Adjusted 2 | | Adjusted 3 | | Adjusted 4 | | Adjusted 5 | |
|---|---|---|---|---|---|---|---|---|---|---|---|---|---|
| Voiding postponement | DSM-5 DE | 1.24 (0.76, 2.03) | 0.392 | 1.21 (0.73, 1.99) | 0.462 | 1.27 (0.76, 2.13) | 0.352 | 1.25 (0.75, 2.09) | 0.387 | 1.20 (0.71, 2.01) | 0.494 | 1.17 (0.70, 1.98) | 0.547 |
| Nocturia | DSM-5 DE | 1.42 (0.80, 2.52) | 0.231 | 1.45 (0.81, 2.60) | 0.213 | 1.42 (0.78, 2.58) | 0.251 | 1.40 (0.77, 2.55) | 0.272 | 1.39 (0.76, 2.54) | 0.282 | 1.31 (0.71, 2.42) | 0.390 |

DSM-5 DE- Disordered eating at Diagnostic and Statistical Manual of Mental Disorders, Fifth Edition frequency (at least once a week). †Analyses performed on n=1494 due to 34 observations being dropped (underweight BMI predicted outcome perfectly). ♦Analyses performed on n=1349 due to 26 observations being dropped (underweight BMI predicted outcome perfectly).

Adjusted 1 confounders: sex. Adjusted 2 confounders: sex and socioeconomic indicators (parental occupational social class, maternal educational attainment, family size, ethnicity, home ownership status, material hardship). Adjusted 3 confounders: sex, socioeconomic indicators, child IQ and developmental level. Adjusted 4 confounders: sex, socioeconomic indicators, child IQ, developmental level, maternal stressful life events, maternal depression and maternal anxiety. Adjusted 5 confounders: sex, socioeconomic indicators, child IQ, developmental level, maternal stressful life events, maternal depression, maternal anxiety, child Body Mass Index and earlier emotional/behaviour problems (fully adjusted model).
